# Addressing the causes of ‘missingness’ in healthcare: a co-designed suite of interventions

**DOI:** 10.1101/2025.10.06.25336469

**Authors:** Calum Lindsay, David Baruffati, Mhairi Mackenzie, David A. Ellis, Michelle Major, Catherine A O’Donnell, Sharon A Simpson, Andrea E Williamson, Geoff Wong

## Abstract

**Background:** Missingness refers to a repeated tendency not to take up offers of care that has a negative impact on the person and their life chances, visible in patterns of missed health appointments. Epidemiological work has shown that patients experiencing ‘missingness’ are more likely to have multiple physical and mental health conditions, to live in adverse or precarious circumstances, and experience a range of negative health outcomes. Yet existing approaches designed to address missed appointments rarely focus on these patients; when they do, it is often through a punitive lens rather than one that engages meaningfully with the causes of missingness. As a result, existing interventions are often ineffective for these patients and may instead worsen access inequalities. This study addresses this gap by outlining a co-produced ‘suite’ of interventions aimed at addressing the specific, complex causes of missingness.

**Methods and Findings:** The study synthesised findings from three co-occurring workstreams: an extensive realist review of 253 documents including peer-reviewed and grey literature; interviews with 61 ‘key informants’ whose personal and professional experiences provided insight into causes and possible solutions; and a series of co-design workshops with a Stakeholder Advisory Group of 16 professionals and experts-by-experience aimed at designing and refining an intervention. The intervention consists of activities across several key domains: embedding a change of perspective around missed appointments; identification; relationships and communication; missingness coordinators; transport and logistics; flexibility; and contact around appointments. Unifying these activities, and crucial to their success, is a paradigm shift towards missed appointments that we term a ‘missingness lens.’

**Conclusions:** This intervention presented here is an evidence-informed, realistic and meaningful set of actions with the potential to address the complex causes of missingness and, by extension, access inequalities and wider health inequalities. Future interventional research into missed appointments should actively focus on or include ‘missing’ patients in design, implementation and the measurement of outcomes.

## Introduction and background

Missed appointments have become central to policy discourse around healthcare globally (1). In multiple country contexts, health services generally – and primary care in particular – are impacted by overlapping crises of growing demand and limited capacity (2). Demand comes through aging populations with increasingly complex health needs, societal mental health crises, and a rise in poverty and inequality alongside the erosion of social safety nets (3, 4). Diminished capacity is caused by chronic underfunding, shortages of appropriate staffing and infrastructure; unsustainable workloads and staff burnout; long waiting lists, and thus services that often appear unresponsive and inaccessible to patients (2, 5, 6). In this context, policymakers often frame missed appointments as a growing source of wasted money and lost capacity, and addressing them becomes a financial and moral – even existential - imperative (7, 8). Much existing research is framed in these terms, as it seeks cost-effective and efficient reductions in missed appointments to benefit services. As outlined previously (9), the issue is often framed as a matter of ‘fixing’ problematic patient behaviours, with theoretical engagements limited to cognitive or behavioural domains, seeking to influence or ‘nudge’ behaviour: reminders; posters or letters seeking to induce commitment guilt or moral responsibility, and the threat sanctions including de-registrations, removal from waiting lists, or fines (10–14).

### The missingness study

Where patients miss *multiple* appointments, the individual behavioural lens often results in overt or covert stigmatising and Othering narratives, with patients labelled as “repeat offenders” (15, 16), “chaotic” or irresponsible (14, 17). The complex health, personal and social circumstances of ‘missing’ patients’ lives, evidenced in preceding epidemiological work (18–21), may also attract stigmatising narratives around poverty, substance use, mental health and long-term health conditions – areas that are also frequently ascribed to failures of individual responsibility (22–24).

Yet as our work has shown, the experience of ‘missingness’ – defined as “the repeated tendency not to take up offers of care that has a negative impact on the person and their life chances” (25) – has a range of complex, overlapping and mutually reinforcing causes that occur at several points throughout patients’ journeys to and through health services (19). These include: the sense that healthcare is not ‘for me’ – not necessary, beneficial, appropriate, or safe; past negative experiences including feelings of stigma, discrimination, neglect and abuses of power, as well as broader issues of trauma and relational adversity; a greater exposure to competing demands and crises, with fewer resources to reduce, mitigate or manage those demands; issues managing gatekeeping and access systems, or the unwritten rules of engagement; issues of travel, transport and the ability to move safely to and through healthcare spaces; and an overall sense of mistrust and distrust (19). Societal and institutional structures play a significant role in sustaining the conditions in which missingness occurs. Stigma influences whether people possess the resources needed to access or negotiate care and patterns their encounters with healthcare and other social institutions (Baruffati et al forthcoming). Care itself is unequally distributed, withheld, and often a poor fit with people’s resources, their experiences of poor health, of their adverse circumstances(26).

These experiences are rarely reflected in the design of interventions because they are absent from the existing evidence base. Focused on service benefits, much of the evidence is not stratified according to who is impacted and in what ways – and who may be missing or absent (27, 28). In some studies, people are excluded by recruitment criteria – for example, those with poor mental health, cognitive impairment, learning disabilities, with insufficient English language skills, without phones, with incomplete practice records, or who are otherwise deemed unsuitable (29–32). Few studies reflect on self-selection bias and those patient groups who may be less likely to accept or use the intervention being tested (33). These absences are structurally patterned, with those excluded often at greater risk of access issues or negative outcomes. Lacking a phone may be a sign of particular adversity (34); the absence of practice data may reflect prior access issues, exclusion, or mis/distrust (27, 35). The limited evidence that does consider patients most likely to be ‘missing’ shows that they are either least likely to benefit from or are actively disadvantaged by interventions designed to ‘work’ for all patients (27, 36, 37). Some consider cancellation as a positive outcome as it reduces the unexpected non-attendance rate for the service (38–40) but this still represents a missed opportunity for care and few studies explore *who* is most likely to cancel, nor whether reductions represent a different form of exclusion as patients are deterred from approaching health services in the first place (27). Thus, throughout the evidence base, patients at risk of missingness also experience “structural missingness” (41), with their situated and experiential knowledge absent from conversations about access to healthcare. The aim of this paper is to address this substantial gap by reporting on a study that synthesised extensive primary and secondary data to design a suite of co-produced, evidence-informed interventions to address missingness in primary care.

## Methods

The intervention development approach was guided by realist principles and by the Six Steps in Quality Intervention Development (6SQuID) approach (42). Here we focus on the first four steps: defining and understanding the problem; identifying malleable factors which have scope for change; identifying how to bring about change (the active ingredient or change mechanism); and exploring how to deliver the change mechanisms. The final two stages involve implementing and evaluating the intervention and are not addressed here. Interventions designed through 6SQuID involve “changing relationships, displacing existing activities and redistributing and transforming resources” (42). They use a systems lens to recentre the role of systemic and contextual causal factors, which are often absent from predominantly behavioural approaches (43).

The stages of the 6SQuID process were addressed through the iterative synthesis of data from the three workstreams of the “developing interventions to reduce ‘missingness’ in health care” project. The study design was approved by the University of Glasgow College of Medical, Veterinary and Life Sciences Ethics Committee in 2022 (Reference 00220187). Workstream 1 was a realist review of literature, identified through database searching and citation-tracking (see (9) for full methods) complemented by further citation tracking and ‘umbrella’ reviews of intervention ideas identified in other workstreams. A full PRISMA diagram and list of included documents is available in sections 1 and 2 of the supporting information. Workstream 2 consisted of realist interviews with 61 key informants whose personal and professional experiences brought a broad range of clinical, social and inclusion health perspectives on missingness. Participant recruitment began on 28^th^ February 2023 and concluded on 11th July 2024. All participants provided informed consent in writing or, where this was not possible, consent was provided verbally and audio-recorded prior to interview. Interviews focused on participants’ theories about the causes of missingness, actions that might address these causes, and factors contributing to or inhibiting their success (see (26) and **Baruffati et al forthcoming** for full methods). Interviews provided further insight into missingness and addressed many of the gaps and shortcomings in the research literature. A breakdown of interview participants is available in section 3 of the supporting information.

The third workstream involved a series of Stakeholder Advisory Group (StAG) workshops. StAG membership was largely drawn from interview participants, with 16 members (8 experts-by-experience and 8 professionals) who reflected a broad range of patient and professional perspectives in the key areas of inclusion health; mental health; homelessness; asylum and migration; substance use; and “severe and multiple disadvantage” (44). The group met regularly throughout the project, acting as guides and sense-checkers for the other workstreams and as co-designers of an overall interventional approach (45). The contribution of each method to the 6SQuID approach is outlined in table 1, and the materials used to guide stakeholder discussions are included in section 6 of the supporting information. Participants in the StAG process provided informed consent in writing to participate in this process.

**Table 1:**
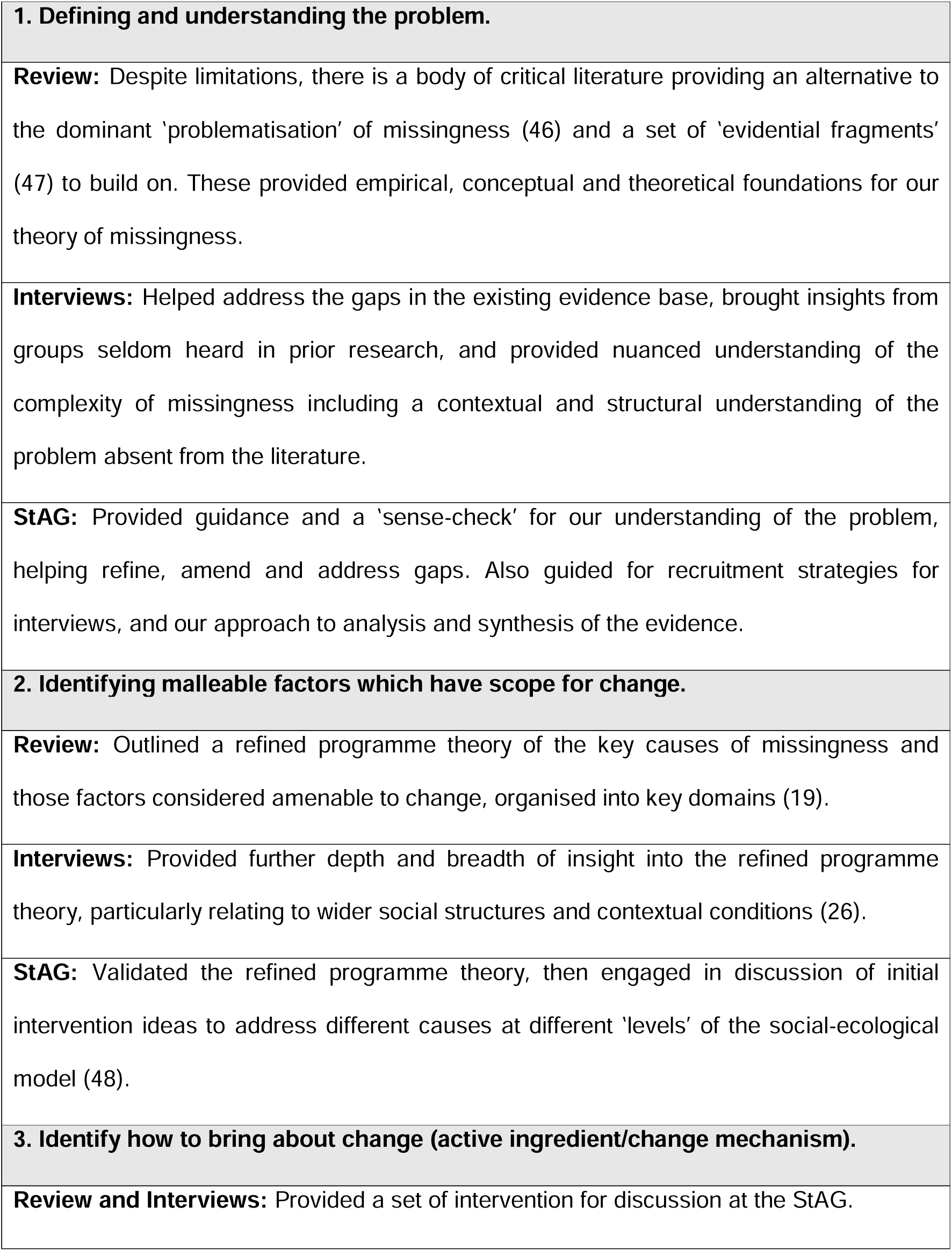
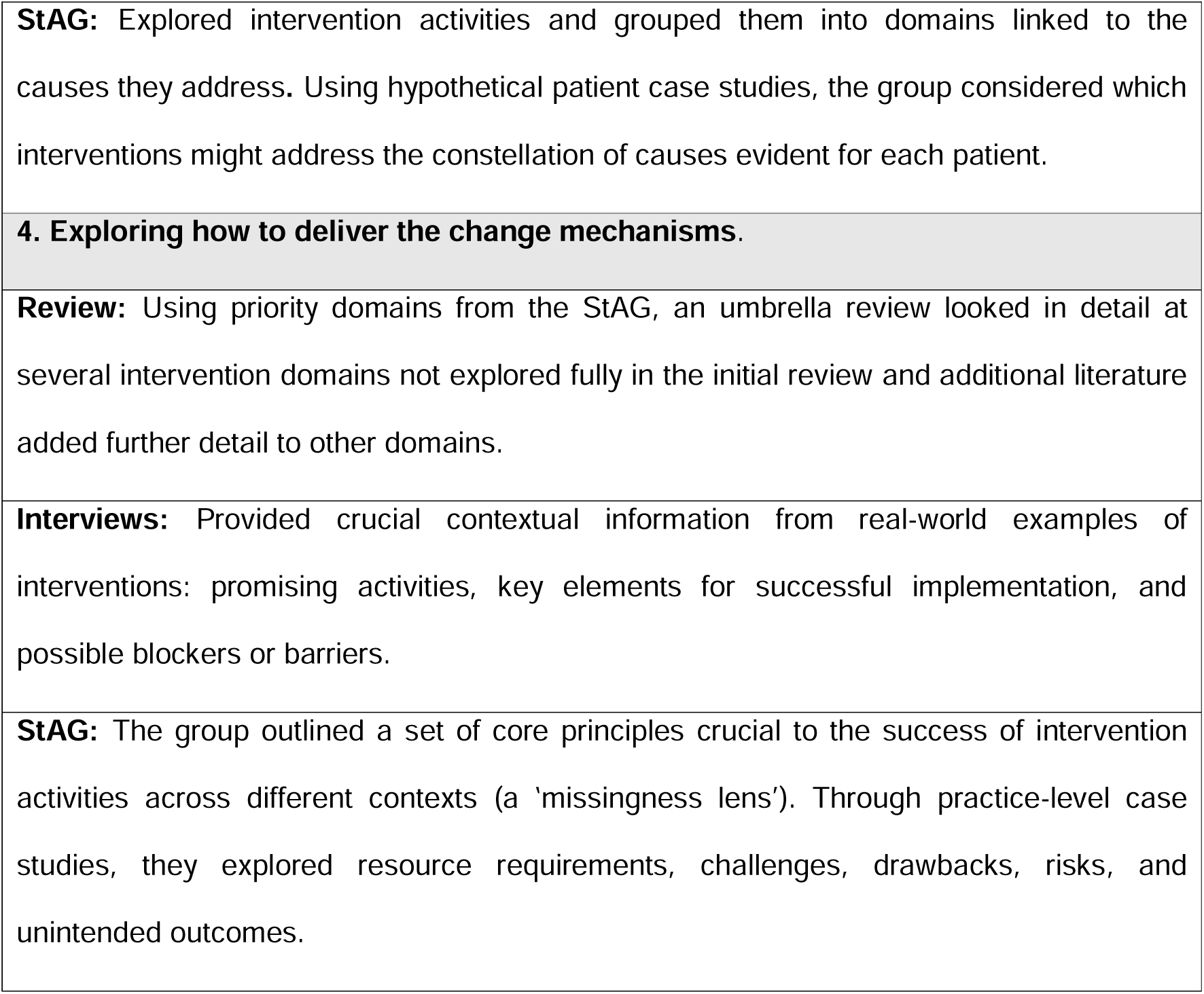
Contribution of workstreams to the 6SQuID process.

Moving back and forth between workstreams allowed each to address the limitations of the others. For example, interventional literature is characterised by significant heterogeneity of settings or approaches to the same activity, *and* by a lack of contextual description, making assessment of implementation processes barriers, risks or unintended outcomes difficult (49–51). These were then explored in interviews and stakeholder workshops. Similarly, the alternative version of the problem evident in interviews and StAGs helped identify additional causes at interpersonal, institutional, community and structural levels, and thus alternative intervention activities for exploration in the iterative literature review process.

The process of analysis was similarly recursive. Literature was analysed in Nvivo, with data relevant to the causal working and contextual dynamics of interventions extracted and synthesised into an explanatory narrative in potential intervention domains. Interviews were analysed separately in Nvivo, coded abductively according to a theoretical framework combining fundamental causation theory and the candidacy framework (see **Baruffati et al (forthcoming)** for further details). Findings from the literature were refined further by findings from interviews, and both used to create materials presented to and refined further by StAG members. Audio recordings and visual materials from StAG meetings were summarised and synthesised with discussions from previous meetings into a final intervention. The final suite of interventions detailed here was synthesised by the research team using summary documents from all three workstreams and was validated by participants at the final StAG. Given the quantity of the data collected, and the nature of the synthesis process above, results are presented as a narrative without primary or secondary data specifically referenced. The contribution of each work package to each intervention domain is described instead in section 7 of the supporting information. Primary data are available in the final project report (forthcoming) and are available on the Open Science Framework (52), congruent with the principles of open research.

## Results

### Key principles: a missingness lens

The intervention presented here is more accurately a ‘suite’ of interventions, or a “recyclable core set of processes” that can be applied according to the requirements of different contexts (53). Their successful implementation depends on creating “congruence” (53) between each part by rooting them within a set of principles termed ‘the missingness lens’ (see figure 1). These principles were defined during a StAG meeting, and they represent a fundamental shift in how the ‘problem’ of missingness is understood and how solutions to it should be designed and implemented.

**Figure 1:**
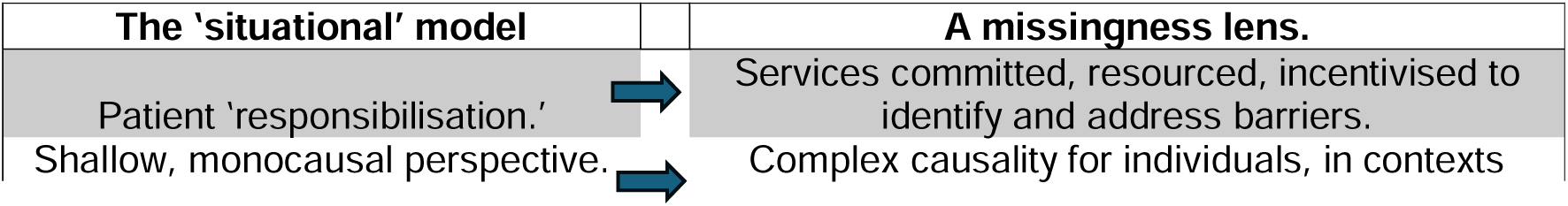
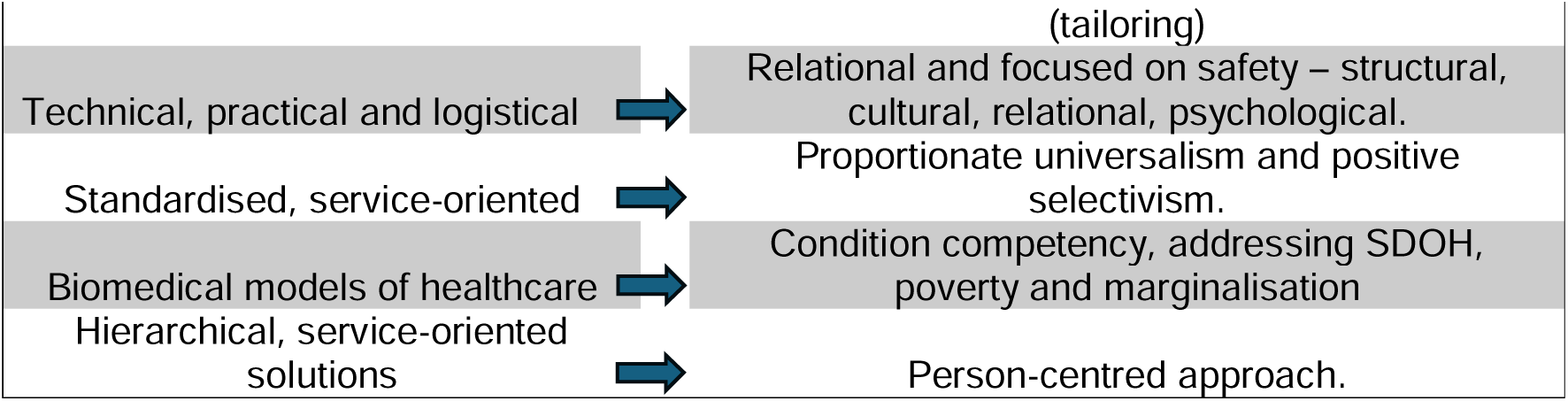
the ‘missingness’ lens.

Firstly, instead of considering missingness as a problem for services caused by patient behaviour, a missingness lens means services commit to directing resources to addressing missingness as a problem for patients and their health. Rather than designing, implementing and measuring interventions across an unstratified patient population, a missingness lens involves both “proportionate universalism” (54)-shifting resources towards those in greatest need - and “positive selectivism” (55) by targeting specific patients and evaluating outcomes for them. It means moving from shallow, simple explanations towards deep and complex causes at multiple levels: structural influences on patients’ circumstances and resources; institutional dynamics of service provision; and interpersonal and intrapersonal processes. It also means moving beyond logistical, technical or practical solutions by recentring affective, psychological and relational elements, particularly around *safety -* psychological safety through trauma-informed practice and psychologically-informed environments (56); and cultural and structural safety by identifying and addressing sources of stigma, discrimination and power imbalance (57). This includes a central role for patient-centred, collaborative care. Finally, a missingness lens moves beyond biomedical and clinical approaches to understand health and healthcare in wider context through condition competency – “clinical understanding of the lived-experience of specific conditions” (26) – and structural competency, the ability to recognise and to influence the social and structural determinants of health (58).

### A ‘suite’ of interventions

The specific suite of actions outlined below represent an attempt to disrupt the systemic causal dynamics sustaining missingness, either directly or by pursuing intermediate outcomes in support of this goal (43, 59). They are limited to those actions which health services have the power to change, while cognisant of the need for broader policy changes to address social determinants of missingness and of illness. The actions are split into several interconnected domains but are likely to be most impactful when synthesised rather than as a set of discrete elements. The domains are represented in figure 2 below, followed by detailed discussion of how each can by implemented in a way that is congruent with the missingness lens.

**Figure 2:**
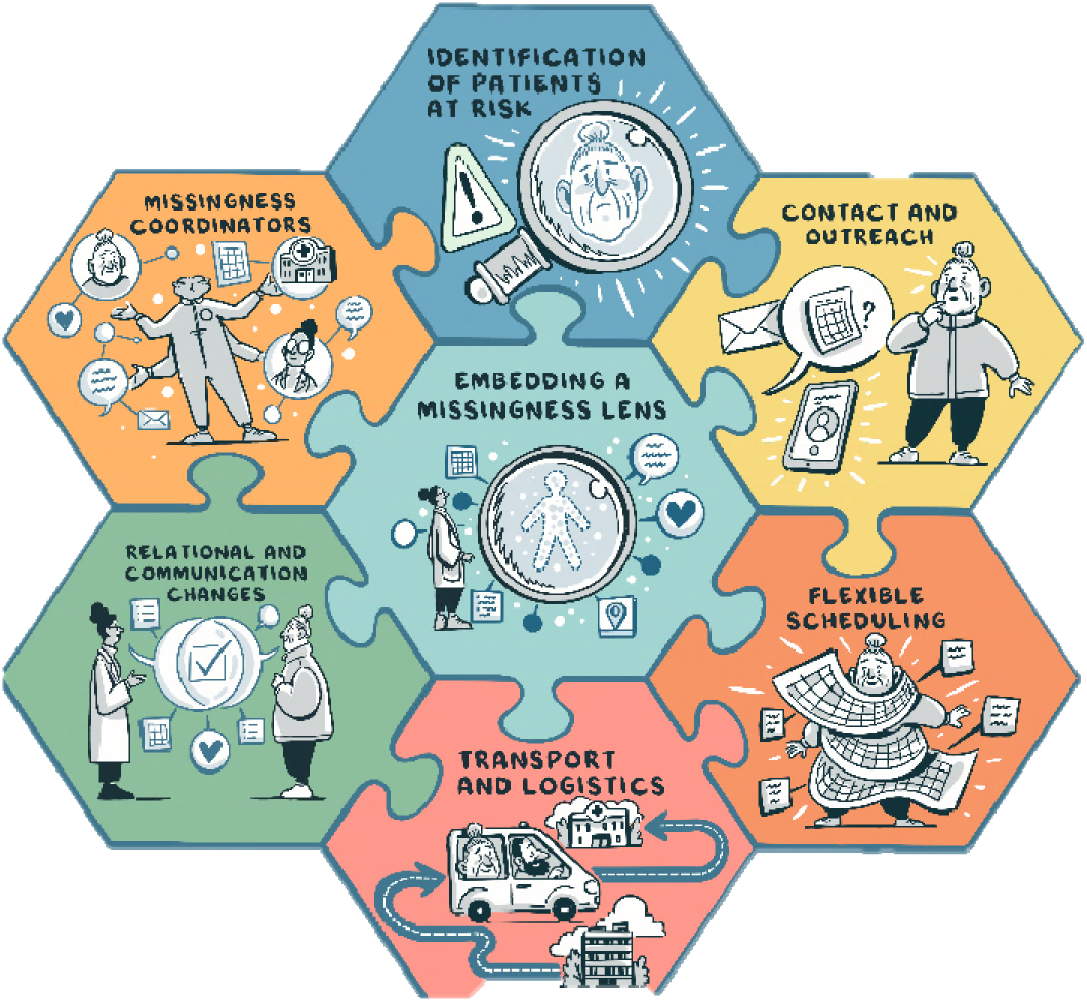
a ‘suite’ of interventions.

### Embedding a missingness lens

The first step in addressing missingness is in embedding the elements of this perspective shift at all levels of health service organisation and delivery. Participants often described the need for a “culture change” towards missed appointments at multiple levels - clinical, administrative, policy, planning, and funding. Resources including money, time, staffing, data infrastructure, knowledge and skills, need to be mobilised and prioritised to ensure this culture change is matched with concrete changes in practices and systems. At the broadest level, this means services having the money to ensure that missingness-focused work – likely to be complex, time- and labour-intensive – is supported, encouraged and even incentivised. Many participants felt that current NHS funding structures like the Quality and Outcomes Framework actively disincentivise missingness-related work, with “exception-reporting” used to exclude patients from monitoring and reporting, rather than incentivising work to get patients into practices. Without specific funding, even the best intentions would remain a “fantasy” or a “dream.” Funding should permit localised innovation and plans for making and monitoring change through multi-component quality improvement methods: setting clear and transparent goals, planning and carrying out key actions, and regularly reviewing or amending them through inclusive, proactive systems for accountability. Bringing together staff and patient perspectives collaboratively is a crucial part of this process. Targeted action, built on solid principles and localised planning, matched with resources (staffing, knowledge, financial), acts as the foundation upon which the activities below can be built. Monitoring and accountability mean going beyond service-side metrics of missed appointment rates to measure impacts on patients qualitatively and quantitatively. They should also occur above individual care delivery, at the strategic and policy level.

Several other actions are suggested to support the embedding process. The first involves training for staff, using knowledge and skills-development to “displace” (42) existing perspectives – particularly stigmatising attitudes or opinions among staff towards missingness or ‘missing’ patients and their health-related or help-seeking behaviours. Training should be provided to all staff at all levels – from administrative to clinical to commissioning – to ensure congruence and consistency. In theory, raising awareness of the negative health outcomes around missingness, of its main causes, and of actionable solutions, may lead to increased staff motivation and empathy, to buy-in for meaningful action, and for patients the sense that services are supportive and safe. All three workstreams suggested other, complementary areas for professional development, including poverty and inclusion health, stigma and discrimination, mental health, psychological trauma, neurodivergence, and attachment. Training should be designed to help staff reflect on practice systems and on relationships and communication dynamics: how and why patients might present differently; how to manage challenging encounters; and how to enact principles of relational care and to create safe environments. Expertise-by-experience is important in formulating and delivering training activities to drive awareness, empathy and change.

However, focusing exclusively on staff attitudes to embed the missingness lens risks simply reversing the direction of blame without acknowledging staff who are motivated to change practice but are inhibited by limited resources and capacity and the burnout they cause. Knowledge, skills and motivation need to be matched by concrete resources to turn motivation into meaningful action. Staff need the protected time and capacity to engage with training and development, and providers the capacity to allow staff to attend as well as the access to training resources and expertise. To allow the missingness lens to displace existing practices, training needs to be complemented by supervision and reflective space, positive reinforcement and feedback, and ongoing support for staff. There is also a role for utilising existing staff knowledge and experience of missingness. Staffing changes may be appropriate, including actively recruiting new clinical and non-clinical staff from communities identified as being at risk of missingness, who may bring knowledge of barriers to care and may act as a bridge to those communities.

### Identification

To build a localised plan for missingness, and to monitor its success, practices and services require IT infrastructure and data recording systems to monitor ‘missingness’ trends and outcomes. Services need systems to identify who is missing through retrospective analysis of appointment trends, and to identify and flag those who become missing in real-time. Past work has demonstrated some difficulties in using UK primary care systems in this way (18, 21, 60). Practices should prioritise the creation of robust coding of patient records, particularly of non-attendance, and should seek to address missing data which is often a site of inequality (41). Services can complement this with other ways to identify those who are missing, for example notifications of ‘missingness’ from referrals to secondary or tertiary care, or staff knowledge of the patient population. While recently ‘machine learning’ approaches have been designed to play this role, caution is needed due to concerns about inaccuracy, bias and the absence of a critical approach to causes in these systems (**Ellis et al, forthcoming).**

In carrying out identification, services must decide on their thresholds for ‘missingness’, particularly considering current resource environments. The initial epidemiological work on missingness selected those with 2 or more missed appointments over a three-year period, based on a pilot study and stakeholder group validation (18, 21, 61). Missingness impacted 19% of the total patient population – a cohort of patients with complex health and social needs (18, 20, 21). Uniformly applying this intervention to all these patients simultaneously risks diluting available resources and limiting impact. To resolve this, services may elect to focus only on those who miss the greatest number of appointments, or whose patient profiles suggested particular vulnerabilities. Crucially, this work is dynamic and recursive and can change as projects develop, and as ideas are tested and approaches refined over time.

Services should complement pattern-identification with identification of the causes of missingness, carried out by contacting those experiencing it and having non-judgemental, respectful and invitational conversations about their experiences. These conversations may have instrumental benefits in identifying individual and collective causal patterns, and may also represent the first step in a change in relationships (42). Staff become more aware and understanding of patients’ circumstances; patients feel a sense of care and effort on the part of services that may previously have seemed uncaring or exclusionary. More broadly, connections with key community stakeholders, community groups or settings may also improve identification of access barriers among specific groups. Within a QI approach, regular monitoring and feedback inputs from staff, individual patients, and across the practice population on areas relevant to missingness acts as a point of ongoing identification.

The findings from identification can be used in two ways. Firstly, they can feed into broader practice changes aimed at improving access to care. Secondly, they can underpin an approach to positive selectivism and proportionate universalism, where particular patients or groups are targeted for tailored responses and additional resources (54, 62). This is closer to individual *needs* identification, covering the specific and proximate causes of missingness in a collaborative and person-centred way, actively seeking and building on patients’ priorities and perspectives, and resulting in a concrete and tailored set of actions from services. A Patient Individual Needs (PIN) document could be created, detailing a patient’s key information and priorities; their access needs and communication preferences; and the steps to be taken by services to support them. Services may also seek insight from significant people or services in patients’ lives (with their consent). The findings of these assessments become the foundations of consistency, continuity and accountability, reducing patients’ needs to advocate for access or additional support, or to repeatedly disclose sensitive information to staff in their pursuit of care. Stakeholders felt that this document should be NHS-wide, ensuring consistency across the system. In line with a person-centred approach these assessments and the support following from them should not be limited to clinical or access issues. Possessing few resources to manage competing demands in multiple domains is a central cause of missingness; if not identified and addressed, these causes of missingness and of ill health are likely to endure (9).

### Relationships and Communication

All workstreams explored the importance of relationships in addressing missingness, and the centrality of trust, consistency, continuity and safety for patients whose service interactions have often been patterned by stigma, discrimination, exclusion, neglect, even hostility. A missingness lens involves recentring the relational causes of missed appointments and including relational components throughout the intervention – for example, the role of training in relational care and trauma-informed, non-stigmatising practice in embedding a missingness lens, or the relational work of reaching out to patients to identify their needs. The outcome of these actions, and the core of a relational approach, is in creating consistency and continuity at every stage of the patient journey, from registration to consultation and follow-up. Patients’ anxieties or fears will likely be reduced if they can trust in a response that is consistently welcoming, invitational, non-judgemental and supportive, not reprimanding, punitive, coercive or manipulative. For those with profoundly negative relational histories, this is even more important. This is not to say that these relationships are unboundaried – instead, clear, agreed boundaries and reciprocal expectations are central to consistency.

Several concrete actions are suggested here. Extending efforts at continuity of care to ‘missing’ patients is central. Being able to see a familiar clinician may reduce anxieties about attendance and increase feelings of safety and security and may also create a sense of reciprocity or loyalty and a relationship to be protected by attending. Being familiar to a clinician or key staff member may encourage “condition competency” (26) and “structural competency” (63) and care plans built upon patients’ needs, circumstances and preferences. Continuity does not have to mean seeing the same person at every interaction. A dedicated small team of clinical and non-clinical within practices focused on missing patients may provide sufficient continuity and retain some flexibility for practices. If patients can contact practices with the knowledge of who they might speak to and how they might be received, by staff aware of their circumstances and needs, anxieties around gatekeeping and initial access may be reduced. Other concrete actions include identifying and catering to patients’ communication needs and preferences in all interactions (e.g language barriers, literacy issues, cognitive challenges, communication preferences).

Relational work takes time. Given the levels of mistrust and past trauma often evident in missingness, people may not instantly respond to relationship-building work. Identification-work will often require a relationship to be established over time, so that the individual feels safe and able to disclose important information about themselves. Persistence is central, but there is also a need to respect patients’ right not be involved and to avoid being overly persistent, coercive or manipulative, or perpetuating problematic power dynamics. A patient-centred and collaborative approach to the intervention and to clinical practice, coupled with a non-judgemental, supportive approach, may contribute to a sense that care is ‘for me’ both instrumentally and relationally.

### Missingness Coordination

The additional work required to address missingness brings the question of *who* is best positioned to do it. Addressing missingness effectively requires a ‘missingness coordinator’, a role akin to other non-clinical roles (e.g navigators, care coordinators, peer workers, mentors, community outreach workers) but oriented specifically around missingness. Missingness coordination involves several key tasks. Firstly, coordinators carry out identification-work and build relationships with patients, seeking to develop trust through active, empathetic, non-judgemental and collaborative support. Coordination is a core part of relational continuity and consistency, a reliable point of contact for the patient and an advocate for them and for good relational practice. In this, missingness coordinators engage in bridging, brokering or mediating activities aimed at creating “safe passage” (64) to and through services. Bridging activities may include working with patients to build their personal resources – confidence, motivation, self-esteem, even expectations for health – as well as supporting changes in service provision through individual, institutional and systemic advocacy, such as the advocacy against austerity carried out by Deep End GPs (65). Care needs to be taken to avoid these workers being made solely responsible for missingness, or their work being marginalised or even co-opted by existing problematic practices. This requires that coordinators be skilled and well-supported, and that they be embedded in services in ways that allow them to change practice in line with the missingness lens.

Using a holistic, person-centred approach to identification, coordination extends beyond primary care settings. It may involve coordinating healthcare across multiple service settings to reduce treatment burden, or brokering access to wider resources aimed at addressing social determinants of illness, social needs or the precariousness that impedes good health and healthcare access. The shape of the work will vary from patient to patient, and thus coordination requires flexibility, open-endedness and a tailored and person-centred approach.

Additional considerations include who is best placed to act as a coordinator, and how they might be funded or positioned to maximise their impact. Expertise-by-experience and ‘peer-ness’ may provide a route to credibility, relationship-building and cultural and structural safety, although stakeholders and interviewees felt that coordinators can come from a range of personal and professional backgrounds as long as they are sufficiently trained, skilled and supported to engage with complex and difficult circumstances. This, plus the need to ‘embed’ workers as equal partners, suggested that paid staff were more appropriate than volunteers. While NHS funding may provide a route to embeddedness, third-sector independence may be a virtue for those mistrustful of the healthcare system.

### Flexibility

Given inflexibility or ‘impermeability’ of services are a central cause of missingness (19), addressing it means changing practice and service systems to be more flexible and suitable for different patient circumstances. There is strong support across all strands of work for prioritising ‘missing’ patients for different forms of tailored flexibility. This includes flexibility in how appointments are made – in person, online, outside the usual appointment-making windows, bypassing reception gatekeeping or telephone triage, booking via coordinators or directly with ‘missingness’-focused staff who can facilitate access. Offering greater choice in when appointments occur may maximise convenience and minimise interference from other demands, as may offering appointments outside of regular hours to those experiencing barriers at other times. Prioritising missing patients for rapid access, drop-in access or open appointment slots may allow providers to minimise delays or take advantage of the “window of opportunity” (27) that exists at the point where care is sought. Offering longer appointments, underpinned by person-centred consultation styles, may have multiple benefits – more time and space to build rapport, for patients to describe needs, circumstances, concerns or preferences, and to discuss and negotiate proposed courses of action. These also become an opportunity to maximise the impact of each single appointment, and thus the perceived benefits of attending in future. Similarly coordinated care planning, or having the flexibility to combine appointments with other health or non-health service providers, may increase the value of attendance and minimise the burdens of managing multiple engagements.

Alongside temporal flexibility, services should offer flexibility in *who* patients see (see relationships and communication above) and in *where* patients are seen. This may mean a role for home visits, or for outreach/in-reach into key settings where identification work suggests ‘missing’ patients are likely to be found (e.g. homelessness services, substance use services, community organisations). In the early stages of missingness work, patients may feel safer and more secure in a familiar environment and their transport and logistical issues may be resolved. Outreach or in-reach may build the initial relational links that lead to more regular engagement or contribute to word-of-mouth and the engagement of other ‘missing’ patients. Remote care through telephone or video appointments is also an option: patients may feel safer or more secure in their home environments; travel costs, time or difficulties may be eliminated, as may competing demands like work or childcare.

Flexibility also means making allowances or accommodations for lateness and avoiding punitive or exclusionary responses to non-standard presentation or continued non-attendance. The shape of flexibility and its limits can be discussed and agreed with patients during initial identification, to ensure both they and service providers have a set of clear expectations and boundaries, agreed and recorded in the PIN. Crucially, it should be offered actively and gladly, neither guarded nor gatekept, as this may demonstrate a level of care and commitment on the part of the service that patients respond to - but only if carried out through a missingness lens without which interventions risk being ineffective or exclusionary. Consider telephone or video appointments, where many of the causes of missingness endure and new causes emerge: a lack of access to technology, limited tech literacy, changing contact details, the costs of data or internet access, complex or unreliable systems, mistrust and concerns about privacy or data security, the lack of safe space, the loss of relational immediacy and limitations on rapport and communication. Carried out with an embedded missingness lens, and designing systems with and for ‘missing’ patients, mean these systems are likely to ‘fit’ patients’ preferences or circumstances. This may include using easily accessible systems, ensuring communication preferences are addressed, and providing support through coordinators to access or use devices or data, connect to systems and facilitate communication. Finally, there is a role for coordinators facilitating flexibility beyond primary care, where ‘missing’ patients might be prioritised to overcome access barriers in secondary and tertiary care including long delays, requirements for re-referral after missed appointments, and exclusionary eligibility criteria.

### Transport and Logistics

As well as being an aspect of flexibility, changing the site or the mode of an appointment is a logistical intervention aimed at reducing barriers related to travel – costs and time, reliance on support networks or unreliable transport systems, limited mobilities, poor mental health, feelings of unsafety (19). There is a spectrum of possible actions here, their suitability dependent on the circumstances of patients and the resources available to practices. At the least intensive end, practices may consider reimbursing travel costs, although this depends on patients having sufficient money to cover costs initially. Services may pre-emptively provide tickets, tokens, vouchers or parking permits instead. Facilitated access to NHS or voluntary transport services may also be beneficial. At the more intensive end, taxi transport and home visits may be most appropriate, as may accompaniment by a missingness coordinator to support safe “wayfinding” (66). Accompaniment can also mean supporting patients within consultations – supporting the exchange of information, providing reassurance or advocacy, and reducing anxieties or worries about attending. Systems for arranging transport need to be accessible, inclusive, and actively offered, as often they are described as inaccessible, complex, inflexible and either actively gatekept or implicitly patterned by eligibility criteria or narratives of deservingness that deter access. Changing service spaces using trauma-informed principles, or setting aside quiet or private waiting rooms, are also crucial to ensuring people feel safe and secure.

### Contact around appointments

Rather than a domain of ‘reminders’, our evidence suggests an expansive approach to patient contact around appointments. Reminding has a role, but reminders alone will not address deeper, broader and more enduring barriers than the simple forgetfulness that most systems aim to address. Basic SMS, automated or email reminders are a minimum standard – the medium determined by patients’ contact preferences, with messages including details of the appointment (time, location, staff member, even purpose). “Stepped” reminders (27) may be beneficial – multiple messages, perhaps sent using different methods, with one reminder far enough in advance to allow patients to plan and other closer to the time to ensure patients do remember. Patients’ preferences should again guide this action. Concerns about data security, privacy and confidentiality should be discussed and addressed where possible – and where patients express a preference to avoid reminders, this should be respected. Practices should prioritise systems for patients to respond to reminders with minimal barriers in order to discuss, cancel or rearrange their appointments. They should work to obtain patients’ contact details and preferences on practice systems, and to maintain these systems to account for changes, otherwise these systems risk perpetuating inequalities. Patients should have the option to identify significant others to contact if they are unavailable.

Yet simply increasing the intensity or quantity of reminders, or tweaking wordings or formats, are unlikely to address deeper causes. There is no support here for behavioural ‘nudges’ in contact around appointments, which were seen to be stigmatising, coercive and alienating. Instead, services should explore personalised pre-appointment contact with identified patients. Personalised contact, carried out by coordinators, may help the relationship-building process with an invitational, welcoming and supportive approach. It may help support orientation as staff and patients discuss what to expect, explore concerns and potential benefits, and identify barriers that are then addressed by actions in other intervention domains.

Contact around appointments also includes contact after appointments, particularly if they are missed, as an opportunity to express care and concern, maintain connection and to check in on the patient’s wellbeing and any health needs or unaddressed access barriers. This should be framed as an invitational offer of support, rather than as a reprimand. The approach to contact should be outlined during identification and agreed in the PIN, so patients know they will be contacted and who will be contacting them, by what method, and with what purpose. Crucially, contact around appointments needs to be two-way. Patients must have the means to respond to written reminders to query or amend details or discuss any concerns. There also needs to be room for patient-initiated contact between or around appointments, whether via clinical teams, dedicated contact lines, or through coordinators, otherwise contact systems risk perpetuating asymmetries where services can freely contact patients but the patient cannot contact them in return.

### Other approaches

Several possible actions are not included here. Some are excluded because stakeholders did not discuss them or because they saw limited value or active drawbacks to using them. Incentivisation of attendance (financial, in kind, or contingency management) did not feature strongly, perhaps reflecting concerns about making healthcare transactional rather than relational, and perpetuating power dynamics and trust. There was no support for fines, punishments or sanctions, or for overbooking approaches that are essentially punitive and were described by one StAG member as “sinister.” Other approaches are promising but fall beyond the purview of individual services – such as ‘hub’ models and integrated services, bringing multiple support services into spaces and thus increasing coordination and reducing the burdens of accessing multiple services. Other aspects are simply beyond the power of health services to change – poverty, austerity, poor housing, the migration system, stigmatising policy discourse, and other determinants of illness – although these continue to act as major impediments to care and to good health. The health service does retain an influencing and advocacy role here, however, as the site where many of the ill effects of broader policy are often most visible.

## Discussion and conclusions

This paper outlines the main components of a major paradigm and practice shift about multiple missed appointments in primary care, based on the application of a missingness lens and of key resources targeted at addressing the complex causes of missingness. The suite of interventions outlined here represent a core set of activities to be applied flexibly according to local contextual factors. A comprehensive approach to missingness would include all the proposed actions, but we recognise the constraints of current primary care systems and the need to prioritise some patients, and some actions, until resourcing permits a more expansive approach.

This study has several strengths. Faced with a literature base built without key perspectives, causal insights or theories of change, our stakeholders and interviewees brought alternative (and seldom-heard) understandings and theories from their direct, contextualised experiences (67). Using knowledge from within systems to build an intervention increases its relevance, practicality and feasibility, and thus its potential for success (42, 68). While there was a high degree of congruence between workstreams, areas of disagreement between data sources could often be accounted for by the flaws of the literature base, or simply by difference of experience, context, preference or positionality between participants. What ‘works’ for one person may not work for another. These differences illuminate, rather than obscure, important causal and contextual dynamics, and highlight the importance of a tailored approach. A further strength relates to the relevance of this work beyond primary care, as both the primary and secondary data include evidence from across the healthcare system.

Although this study is evidence-informed and based on thorough theorising, it does not test or evaluate the proposed intervention in practice. However, it does provide some implications for future research and evaluation of interventions into missingness and missed appointments more generally. Some we have detailed previously – the need to change the framing of missed appointments, to include data and analysis on missingness, and to use inclusive methods of recruitment and data collection (19). Ensuring data is equally available for ‘missing’ patients, acknowledging where and why it is not, and stratifying analysis of intervention effects according to how they impact those experiencing missingness, are crucial ways to counter “structural missingness” (41). Interventional research should also carefully consider what outcomes are measured, with a stronger focus on patient health and wellbeing and on patients’ *chosen* outcomes in a person-centred approach. It should not only measure numerical reductions in missed appointments, or in missingness, but should also explore intermediate outcomes related to causes: do patients feel cared for, supported and understood? Is their experience of care or its quality improved? Does their health or quality of life improve? Crucially, how are these improvements unequally distributed? Finally, greater reporting of the context and process of interventions is crucial to improving an evidence base where these insights are limited.

A further limitation of this study relates to its ‘level’ of focus. During intervention design, contributors considered interactions within and between multiple levels of focus – structural, community, institutional, individual – before focusing on changes within the power of health services. Consequently, many causal factors fall beyond our remit, particularly structural factors: growing poverty and inequality, austerity politics, crumbling transport infrastructure, the decimation of the third sector safety net; hostile immigration policies, and the systematic underfunding of health and social services that erode their capacity and erode public trust within them. If these issues continue to worsen there is a risk that institutional and practice changes become “fantasy paradigms” unable to mitigate deeper and more powerful forces (69).

The suite of interventions outlined here are highly congruent with several recent papers on addressing access and health inequalities in the UK (62, 70–72). Beyond specific Inclusion Health settings such as homelessness health care there are already encouraging mainstream care examples, such as the Inclusion Health Action in General Practice (HAGP) approach in Scotland. ‘Deep End’ practices were resourced to address inequalities, with missingness chosen as a point for action, and were able to evidence improved access and improved health, better relational environments, and improved staff morale and support (60). The unique contribution of this work is that missingness provides a tangible and measurable focal point for work to address both access inequalities and broader health inequalities. These problems are often framed as being particularly complex and intractable, overwhelming and demoralising to those seeking to address them. Addressing missingness provides an anchor point, a framework and a site for meaningful change, and with the application of resources to the problem evidence suggests that profound transformation can be achieved (60, 64, 73–75).

## Data Availability

Yes - all data will be fully available without restriction via the Open Science Framework

https://osf.io/e4bdv/

## Acknowledgements

The research team would like to acknowledge the invaluable contribution of interview participants and members of our Stakeholder Advisory Group to this work. In addition Elspeth Rae, University of Glasgow, provided administrative support; Claire Duddy, formerly university of Oxford, was the information specialised who devised and conducted initial literature searches; and Jack Brougham, illustrator, provided illustrations used in the paper and throughout the project.

