## Supporting information file for "Addressing the causes of ‘missingness’ in healthcare: a co-designed suite of interventions"

### PRISMA diagram

**Identification of studies: Initial Search**

Duplicate records removed from combining database and citation records (n=84)

Duplicate records removed in Distiller (n=4)

Duplicate records removed from databases (n=692)

Records identified from Databases (n=1462)

**Remaining papers:1,372**

Duplicate records removed from citations (n=189)

Records identified from citations from/of existing reviews (n=879)

**Identification**

Records identified from other sources:

colleagues, grey literature, NHS documents, (n=101)

citation alerts for core papers (n = 14)

**Documents remaining for title and abstract screening – 1487**

Records excluded with reasons: (n=1210)

NHS but not primary care (inc. secondary, outpatient, screening etc) (n = 193)

Missed appointments but non-NHS primary care (other countries) (n = 108)

Missed appointments in non-health services, UK (n = 1)

Missed appointments, other health services, non-UK/location unclear (n = 465)

None of the above/not relevant (n = 443)

**Records included: 277**

Meets all criteria: (n = 127)

Unclear/other reasons for full-text inclusion (n = 150)

**Title and abstract screening**

Records included (n = 277)

Texts not retrieved: (n = 2)

**Full texts reviewed (n = 275)**

**Documents included (n=85)**

Excluded, with labels:

No – moved to background (n = 82)

No – relabelled, excluded (n = 104)

Duplicates removed: (n = 4)

**Full-text screening**

**Identification of studies: revisiting excluded papers**

**UK, non-primary care:**

- papers excluded (n=208)
- protocols/posters removed (n=2) and replaced by related peer-reviewed studies (n=4 added).

**UK, non-primary care papers included: (n=42)**

Non-UK, primary care papers excluded (n=99)

**Non-UK, primary care papers included (n=37)**

**Documents included: 79**

Papers about non-attendance (UK, non-primary care) (n=248)

Papers about non-attendance (primary care, non-UK): (n=136)

**Initial of studies: Other sources**

Additional grey literature: **5**

Additional papers from ‘pearling’/found opportunistically: **15**

Citation alerts: **13**

**Total number from initial review: 197**

**Documents included: 33**

Documents included: 56

**Iterative identification of studies: Intervention-specific papers**

**Total number of studies: 253**

Papers from retrospective citation tracking and ongoing citation alerts of key papers/terms: **29**

Papers identified through searching for reviews of key activities: **20**

Additional papers found opportunistically: **7**

### 1. Study characteristics

| **Location** | **No. of studies** | **Health setting** | **No. of studies** |
| --- | --- | --- | --- |
| Australia | 3 | Primary care | 99 |
| Canada | 6 | Outpatient/specialist – physical health | 58 |
| Denmark | 3 | Mental health/substance use | 19 |
| Germany | 2 |  |  |
| Israel | 1 | Healthcare general | 24 |
| Malaysia | 1 | Multiple settings | 15 |
| n.a/other | 56 | Other/n.a/not stated | 38 |
| New Zealand | 1 |  |  |
| Switzerland | 1 |  |  |
| UK | 124 |  |  |
| USA | 55 |  |  |
| **Study design/methods** |  |  |  |
| Qualitative (interview, focus group, observation, or combination of these) | 38 |  |  |
| Systematic review, realist synthesis or meta-analysis | 31 |  |  |
| Literature review, non-systematic (inc. scoping) | 24 |  |  |
| Quantitative (administrative data inc. retrospective, observational, linkage studies) | 57 |  |  |
| Questionnaire/survey | 28 |  |  |
| Mixed (administrative data + questionnaire/survey) | 23 |  |  |
| Mixed (administrative + qualitative data) | 12 |  |  |
| Mixed (other) | 10 |  |  |
| Randomized trials | 6 |  |  |
| Theoretical/conceptual papers | 9 |  |  |
| Policy/guidance documents | 3 |  |  |
| Opinion piece/news article/press releases/letters | 12 |  |  |

### 2. Characteristics of Interview Participants

| **Category** | **Service/Vulnerability** | **No. Participants** |
| --- | --- | --- |
| Professional | Third-Sector Specialist Service pertinent to Inclusion Health^1^ | 10^2^ |
| Professional | NHS Inclusion Health | 8 |
| Professional | Local/National (Scotland/UK) Level Strategic/Policy Role | 6 |
| Professional | NHS Mainstream Healthcare | 5 |
| Professional | NHS Specialist Service pertinent to Inclusion Health | 4 |
| Expert-by-Experience | Participants with experiences conferring risk of missingness: homelessness, the asylum system, poverty, domestic abuse, diabetes, a learning disability; mental health condition(s), problem substance use, having been in care or having a caring role. | 28 |
| Total |  | 61 |

^1^ Inclusion health refers to healthcare approaches targeted at addressing those most negatively affected by health and social inequalities. The term is typically used to refer to those experiencing acute forms of exclusion and marginalisation, such as those experiencing homelessness, prison, and/or problem substance use (Public Health England, 2021).

^2^ This includes three participants who were professionals with personal lived experience of missingness.

Table taken from Baruffati et al **forthcoming.**

### 3. List of included documents

| **Author** | **Title** | **year** | **Source** | **Setting + Geographical area** |
| --- | --- | --- | --- | --- |
| Abdulkadir, L.S.; Mottelson, I.N.; Nielsen, Dorthe, S. | Why does the patient not show up? Clinical case studies in a Danish migrant health clinic | 2019 | Eur J Pers Cent Healthc. 2019;7(2):316-24. | Denmark (migrant health clinic) |
| Adepoju, O.E. | Longer appointment duration reduces future missed appointments in safety-net clinics | 2025 | The American Journal of Managed Care | USA – Safety-Net clinics |
| Aggarwal, A., Davies, J., Sullivan, R. | "Nudge" and the epidemic of missed appointments Can behavioural policies provide a solution for missed appointments in the health service? | 2016 | Journal of Health Organization and Management, 30(4) | n/a (review paper) |
| Åkerblom, K.B. and Ness, O. | Peer workers in co-production and co-creation in mental health and substance use services: a scoping review. | 2023 | Administration and Policy in Mental Health and Mental Health Services Research, 50(2) | Review paper - Substance use services |
| Akter, S., Doran, F., Avila, C., & Nancarrow, S. | A qualitative study of staff perspectives of patient non-attendance in a regional primary healthcare setting | 2014 | Australas Med J. 2014;7(5):218-26. | Australia (primary care) |
| Ali-Faisal, S.F., Colella, T.J., Medina-Jaudes, N. and Scott, L.B. | The effectiveness of patient navigation to improve healthcare utilization outcomes: A meta-analysis of randomized controlled trials | 2017 | Patient Education and Counseling 100 (2017) 436–448 | n/a – review paper. |
| Amberger, C & Schreyer, D. | What do we know about no-show behavior? A systematic, interdisciplinary literature review | 2022 | J Econ Surv. 2022;38:57–96 | n/a (review paper) |
| Anderson, A. | Interventions to reduce did not attend rates for priority (vulnerable) populations | 2025 | Monash Health Library | Various – various |
| Anyaegbu, C.T. | SMS reminders: reducing DNA at a community mental health depot clinic. | 2021 | Journal of Community Nursing, 35(1). | UK (outpatient - community mental health) |
| Arber, S. & Sawyer, L. | Do appointment systems work | 1982 | BMJ. 1982;284(6314):478-80. | UK (primary care) |
| Arnold, O.F. | Reconsidering the “NO SHOW” Stamp: Increasing Cultural Safety by Making Peace with a Colonial Legacy | 2012 | Northern Review (26) | Canada (primary care) |
| Aspinall, P.J. | Inclusive practice: vulnerable migrants, gypsies and travellers, people who are homeless, and sex workers: a review and synthesis of interventions/service models that improve access to primary care & reduce risk of avoidable admission to hospital | 2014 | https://assets.publishing.service.gov.uk/media/5a7db8b2e5274a5eaea65ee4/Inclusive_Practice.pdf | UK (primary care) |
| Ayalde, J., Soong, W., Thomas, S., McCann, P., Griffiths, J., Nicholls, C., Heble, S., Dragovic, M. & Waters, F. | Reasons for non-attendance in youth mental health clinics: Insights from mobile messaging communications | 2022 | Early Interv Psychiatry, 2023;17(9):877-83. | Australia (outpatient – mental health) |
| Ballantyne, M. Liscumb, L., Brandon, E., Jaffar, A., Macdonald, A., Beaune, L. | Mothers’ Perceived Barriers to and Recommendations for Health Care Appointment Keeping for Children Who Have Cerebral Palsy | 2019 | Glob Qual Nurs Res. 2019;6. | Canada (primary care) |
| Bansal, I., Soni, R., Eisen, S., Ward, A., Longley, N., & Sen, C. | Breaking the Barriers to accessing care: co-creating solutions with refugee service users | 2023 | Arch Dis Child. 2023;108. | UK (primary care) |
| Barker, I., Steventon, A., Williamson, R. & Deeny, S.R. | Self-management capability in patients with long-term conditions is associated with reduced healthcare utilisation across a whole health economy: cross-sectional analysis of electronic health records | 2018 | BMJ Qual Saf. 2018;27(12):989-99. | UK (primary care) |
| Barmore, P., Webb, E., Venable, K., Burnette, A., Echols, J.D., Schech, S., Cartie, R. and Sood, R. | A Quality Improvement Project to Impact Adult and Pediatric No-Show Rates. | 2024 | Journal of Burn Care & Research, 45(Supplement_1) | USA – wound and burn clinic |
| Barron, W.M | Failed appointments - who misses them, why they are missed, and what can be done | 1980 | Prim Care. 1980;7(4):563-74. | n/a (review paper) |
| Bean, A.G. & Talaga, J. | Appointment breaking: causes and solutions | 1992 | J Health Care Mark. 1992;12(4):14-25. | n/a (review paper) |
| Bech, M. | The economics of non-attendance and the expected effect of charging a fine on non-attendees | 2005 | Health Policy, 74(2) | Denmark (health general) |
| Bell, J., Gottlieb, L.M., Lyles, C.R., Nguyen, O.K., Ackerman, S.L. and De Marchis, E.H. | Provision of digital devices and internet connectivity to improve synchronous telemedicine access in the US: a systematic scoping review. | 2024 | Frontiers in Digital Health, 6, p.1408170. | USA – review paper. |
| Bickler, C. B. | Defaulted appointments in general practice | 1985 | J R Coll Gen Pract. 985;35(270):19-22. | UK (primary care) |
| Bidmead, E., Hayes, L., Mazzoli-Smith, L., Wildman, J., Rankin, J., Leggott, E., Todd, L. and Bramhall, L. | Poverty proofing healthcare: A qualitative study of barriers to accessing healthcare for low-income families with children in northern England. | 2024 | PLOS One, 19(4), p.e0292983 | UK (England) – healthcare general. |
| Biggs, J., Njoku, N., Kurtz, K., Omar, A. | Decreasing Missed Appointments at a Community Health Center: A Community Collaborative Project | 2022 | J Prim Care Community Health. 2022;13:3. | USA (primary care) |
| Birkenmaier, J., Maynard, B.R., Blumhagen, H.M. and Shanks, H. | Medical‐financial partnerships for improving financial and medical outcomes for lower‐income Americans: A systematic review. | 2024 | Campbell Systematic Reviews, 20(4), p.e70008. | USA – review paper |
| Blankenstein, R. | Failed appointments - Do telephone reminders always work? | 2003 | Clin Gov. 2003;8(3):208-12. | UK (primary care - dental) |
| Boksmati, N., Butler-Henderson, K., Anderson, K., & Sahama, T. | The Effectiveness of SMS Reminders on Appointment Attendance: a Meta-Analysis | 2016 | J Med Syst. 2016;40(4):10. | n/a (review paper) |
| Boos, E.M., Bittner, M.J. & Kramer, M.R. | A Profile of Patients Who Fail to Keep Appointments in a Veterans Affairs Primary Care Clinic | 2016 | WMJ. 2016;115(4):185-90. | USA (primary care - veterans affairs) |
| Boshers, E. B., Cooley, M. E. & Stahnke, B. | Examining no-show rates in a community health centre in the United States | 2021 | Health Soc Care Community. 2022;30(5):e2041-e9. | USA (primary care) |
| Bowser, D.M., Utz, S.G, Doris,Harmon, Rebecca | A Systematic Review of the Relationship of Diabetes Mellitus, Depression, and Missed Appointments in a Low-Income Uninsured Population | 2010 | Arch Psychiatr Nurs. 2010;24(5):317-29. | n/a |
| Brewster, S. | The role of community pharmacy in supporting people with diabetes who have a history of repeated non-attendance at healthcare appointments | 2023 | Doctoral dissertation, University of Southampton | UK (primary care - pharmacy - diabetes) |
| Brown, Sarah | Qualitative evaluation of Focused Care | 2019 | https://focusedcare.org.uk/wp-content/uploads/2020/11/FC-Qualitative-Eval.pdf | UK (primary care) |
| Budde, H., Williams, G.A., Winkelmann, J., Pfirter, L. and Maier, C.B. | The role of patient navigators in ambulatory care: overview of systematic reviews. | 2021 | BMC health services research, 21, pp.1-12. | n/a – review paper |
| Buetow, S. | Non-attendance for health care: When rational beliefs collide | 2007 | Sociol Rev. 2007;55(3):592-610. | n/a |
| Bull, S.L., Frost, Nicki,Bull, Eleanor R. | Behaviourally informed, patient-led interventions to reduce missed appointments in general practice: a 12-month implementation study | 2022 | Fam Pract. 40(1) | UK (primary care) |
| Cameron, E. | A mixed methods investigation of parental factors in non-attendance at general paediatric hospital outpatient appointments | 2015 | Doctoral dissertation, Aston University | UK (primary care + outpatient - paediatrics) |
| Cameron, E., Heath, G., Redwood, S., Greenfield, S., Cummins, C., Kelly, D., Pattison, H. | Health care professionals' views of paediatric outpatient non-attendance: implications for general practice | 2014 | Fam Pract. 2014;31(1):111-7. | UK (primary care + outpatient) |
| Campbell, K., A. Millard, G. McCartney, and S. McCullough. | Who is least likely to attend? An analysis of outpatient appointment ‘Did Not Attend’ (DNA) data in Scotland | 2015 | https://www.healthscotland.scot/media/1129/5348_dna-analysis_nhs-ggc.pdf | UK (outpatient - various) |
| Campbell-Richards, D. | Exploring diabetes non-attendance: An Inner London perspective | 2016 | J Diabetes Nurs. 2016;20(2):73-8. | UK (outpatient - diabetes) |
| Car, J., Sheikh, A. | Telephone consultations | 2003 | BMJ. 2003;326(7396):966-9. | n/a (review paper) |
| Car, J., Ng, C., Atun, R., Card, A. | SMS Text Message Healthcare Appointment Reminders in England | 2008 | J Ambul Care Manage. 2008;31(3):216-9. | UK (health general) |
| Carmichael, C., Smith, L., Aldasoro, E., Gil Salmerón, A., Alhambra-Borrás, T., Doñate-Martínez, A., Seiler-Ramadas, R. and Grabovac, I. | Exploring the application of the navigation model with people experiencing homelessness: a scoping review | 2023 | Journal of Social Distress and Homelessness, 32:2, 352-366, | n/a – review paper – homelessness |
| Carrillo de Albornoz, S., Sia, K.L. and Harris, A. | The effectiveness of teleconsultations in primary care: systematic review | 2022 | Family Practice, 39(1), pp.168-182. | N/a – review paper – primary care/telehealth |
| Carruthers, E., Dobbin, J., Fagan, L., Humphrey, A., Nagasivam, A., Stevenson, K., Yuan, J.M., Aldridge, R.W. and Burns, R. | Interventions to improve access to primary care for inclusion health groups in England: a scoping review | 2023 | The Lancet, 402, p.S32 | UK (England) – primary care – inclusion health |
| Cashman, S. B., Savageau, J. A., Lemay, C. A., Ferguson, W. | Patient Health Status and Appointment Keeping in an Urban Community Health Center | 2004 | J Health Care Poor Underserved. 2004;15(3):474-88. | USA (primary care) |
| Chapman, K.A., Machado, S.S., van der Merwe, K., Bryson, A., Smith, D. | Exploring Primary Care Non-Attendance: A Study of Low-Income Patients | 2022 | J Prim Care Community Health. 2022;13. | USA (primary care) |
| Chen, K., Zhang, C., Gurley, A., Jackson, H., Akkem, S. | Appointment Non-attendance for Telehealth Versus In-Person Primary Care Visits at a Large Public Healthcare System | 2023 | J Gen Intern Med. 2023 Mar;38(4):922-8. | USA (primary care) |
| Ciechanowski, P., Russo, J., Katon, W., Simon, G., Ludman, E., Von Korff, M., Young, B., Lin, E. | Where is the patient? The association of psychosocial factors and missed primary care appointments in patients with diabetes | 2006 | Gen Hosp Psychiatry. 2006;28(1):9-17 | USA (primary care) |
| Clarke, R | An Exploration into how Community Navigation Programmes Support Wellbeing: A Realist Evaluation | 2022 | Doctoral dissertation, University of Lincoln. | UK (England) – community navigation service |
| Claveau, J., Authier, M., Rodrigues, I., Crevier-Tousignant, M. | Patients’ missed appointments in academic family practices in Quebec | 2020 | Can Fam Physician. 2020;66(5):349-55. | Canada (primary care) |
| Corfield, L., Schizas, A., Williams, A., Noorani, A. | Non-attendance at the colorectal clinic: a prospective audit | 2008 | Ann R Coll Surg Engl. 2008;90(5):377-80 | UK (outpatient - colorectal clinic) |
| Corrigan, P.W., Pickett, S., Schmidt, A., Stellon, E., Hantke, E., Kraus, D., Dubke, R. | Peer navigators to promote engagement of homeless African Americans with serious mental illness in primary care | 2017 | Psychiatry Res. 2017;255:101-3. | USA (primary care) |
| Corrigan, P.W., Talluri, S.S. and Shah, B. | Formal peer-support services that address priorities of people with psychiatric disabilities: A systematic review. | 2022 | American Psychologist, 77(9), p.1104. | n/a – peer support – mental health/psychiatry |
| Cosgrove, M.P. | Defaulters in general practice: reasons for default and patterns of attendance | 1990 | Br J Gen Pract. 1990;40(331):50-2. | UK (primary care) |
| Coulter, A., Roberts, S., Dixon, A. | Delivering better services for people with long-term conditions: Building the house of care | 2013 | https://www.kingsfund.org.uk/insight-and-analysis/reports/better-services-people-long-term-conditions | n/a (review paper) |
| Cousins, C., Baxter, J., Javier Vilar, S. R. F. | Non-attendance at hospital clinics for Hepatitis C among intravenous drug users: barriers and potential solutions | 2011 | J Infect. 2011 Dec 1;63(6):e65-6. | UK (outpatient - hepatitis C) |
| Cowling, A., Nevens, L., Contogiorgi, N., Bradley, N., Wilcockson, C., Stebbings, C., McMeekin, P., Steer, M. and Harland, J. | Developing and piloting an intervention to reduce colposcopy non-attendance and corresponding inequalities: a mixed-methods study |  | The Lancet, 402, p.S35. | UK (England) – colposcopy |
| Crable, E.L., Biancarelli, D.L., Aurora, M., Drainoni, M.L. and Walkey, A.J., | Interventions to increase appointment attendance in safety net health centers: A systematic review and meta‐analysis. | 2021 | Journal of Evaluation in Clinical Practice, 27(4), pp.965-975. | USA – review paper – safety net settings |
| Crane, M.A., Cetrano, G., Joly, L.M.A, Coward, S., Daly, B.J.M., Ford, C., Gage, H., Manthorpe, J., Williams, P. | Mapping of specialist primary health care services in England for people who are homeless | 2018 | https://kclpure.kcl.ac.uk/ws/portalfiles/portal/88024144/HEARTH_study_Mapping_SummaryReport_2018.pdf | UK (primary care) |
| Crocker, C., Teehan, M., Ursuliak, Z., Morrison, J., Robertson, N., Alexiadis, M., Tibbo, P. | Patient engagement to early intervention in psychosis services: retrospective analysis of engagement patterns | 2020 | Schizophrenia bulletin, 46(Supplement_1), pp.S129-S130. | Canada (outpatient - mental health) |
| Cummings, C., Raja, P., Gabrielian, S. and Doran, N. | Impacts of Telehealth Adoption on the Quality of Care for Individuals With Serious Mental Illness: Retrospective Observational Analysis of Veterans Affairs Administrative Data. | 2024 | JMIR Mental Health, 11(1), p.e56886. | USA – veterans affairs/mental health |
| Dantas, L. F., Fleck, J.L., Cyrino O., Fernando L., Hamacher, S. | No-shows in appointment scheduling - a systematic literature review | 2018 | Health Policy. 2018;122(4):412-21. | n/a (review paper) |
| Denneny, E. K., Black, S. E., Bogle, Y., Macavei, V. M., O'Shaughnessy, T. C., White, V. L. C., Kunst, H., Jayasekera, N. P. | Tackling poor attendance to tuberculosis clinic – who, why and what can be done | 2014 | Thorax 2014;69:A210. | UK (outpatient - tuberculosis clinic) |
| Dent H., Abdalla H., Segal, S., Osakwe, E. Davies, A.M. | 6577 No child left behind – AI’s role in equalising healthcare | 2024 | Archives of Disease in Childhood 2024;109:A398. | UK (England) – paediatrics. |
| Department for Levelling Up, Housing and Communities | Frontline support models for people experiencing multiple disadvantage: A Rapid Evidence Assessment | 2023 | https://assets.publishing.service.gov.uk/media/642af3507de82b000c31350c/Changing_Futures_Evaluation_-_Frontline_support_models_REA.pdf | Various (literature review) |
| Deyo, R. A., Inui, T. S. | Dropouts and broken appointments. A literature review and agenda for future research | 1980 | Med Care. 1980;18(11):1146-57. | n/a (review paper) |
| Dinsdale, P. | Practice nurses reject fines for missed appointments | 2001 | Nurs Stand (through 2013) 2001 Aug;15(49):8. | UK (primary care) |
| Dockery, F., Rajkumar, C., Chapman, C., Bulpitt, C., Nicholl, C. | The effect of reminder calls in reducing non-attendance rates at care of the elderly clinics | 2001 | Postgrad Med J. 2001;77(903):37-9. | UK (outpatient - care for the elderly) |
| Doraiswamy, S., Jithesh, A., Mamtani, R., Abraham, A., Cheema, S. | Telehealth Use in Geriatrics Care during the COVID-19 Pandemic—A Scoping Review and Evidence Synthesis | 2021 | Int. J. Environ. Res. Public Health 2021, 18, 1755. | Review paper – telehealth/geriatric care. |
| DuMontier, C., Rindfleisch, K., Pruszynski, J., Frey, J.J., | A Multi-Method Intervention to Reduce No-Shows in an Urban Residency Clinic | 2013 | Fam Med. 2013;45(9):634-41 | USA (primary care) |
| Dunmore, C., Baldwin, L., Akpan, A. | Social determinants and older people hospital outpatient non-attendance | 2017 | Age Ageing. 2017;46(Supplement 3):iii1 | UK (outpatient - care for the elderly) |
| Dyer, B. T., Swann, F., Kadam, M., Draper, J., Mc Gill, L. A., Kapetanakis, S., Ismail, T., Carr-White, G., Webb, J. | Understanding non-attendance to an inner city tertiary centre heart failure clinic: a pilot project | 2019 | Eur Heart J. 2019;40(Supplement 1):3747 | UK (outpatient - tertiary heart clinic) |
| Eades, C., Alexander, H. | A mixed‐methods exploration of non‐attendance at diabetes appointments using peer researchers | 2019 | Health Expect. 2019;22(6):1260-71 | UK (outpatient - diabetes) |
| Edwards, K. | Identifying Patient Preferences in Appointment Reminders for Adults to Reduce Missed Appointments in an Outpatient Mental Health Clinic: A Quality Improvement Project | 2023 | Masters dissertation, Georgia State University | USA (outpatient - mental health) |
| Ellis, D. A., Jenkins, R. | Weekday Affects Attendance Rate for Medical Appointments: Large-Scale Data Analysis and Implications | 2012 | PLoS One. 2012;7(12):4. | UK (primary care + outpatient) |
| Ellis, D. A., McQueenie, R., McConnachie, A., Wilson, P., Williamson, A. E. | Demographic and practice factors predicting repeated non-attendance in primary care: a national retrospective cohort analysis | 2017 | Lancet Public Health. 2017;2(12):E551-E9. | UK (primary care) |
| Fairhurst, K., Sheikh, A. | Texting appointment reminders to repeated non-attenders in primary care: randomised controlled study | 2008 | Qual Saf Health Care. 2008;17(5):373-6. | UK (primary care) |
| Fee, P. A., Hargan, A. M. | An intervention study to assess the effectiveness of a reminder telephone call in improving patient appointment attendance at a Community Dental Service clinic | 2016 | Community Dent Health. 2016;33(4):239-41 | UK (primary care - dental) |
| Fernandes, L.G., Devan, H., Fioratti, I., Kamper, S.J., Williams, C.M. and Saragiotto, B.T. | At my own pace, space, and place: a systematic review of qualitative studies of enablers and barriers to telehealth interventions for people with chronic pain. | 2022 | Pain, 163(2), pp.e165-e181. | Review paper – chronic pain/telehealth |
| Fien, S., Dowsett, C., Hunter, C.L., Myooran, J., Sahay, A., Menzel, K. and Cardona, M. | Feasibility, satisfaction, acceptability and safety of telehealth for First Nations and culturally and linguistically diverse people: a scoping review. | 2022 | Public health, 207, pp.119-126. | Review paper – telehealth. |
| Finlayson, S., Boelman, V., Young, R., Kwan, A. | Saving lives, saving money: how homeless health peer advocacy reduces health inequalities | 2015 | https://groundswell.org.uk/wp-content/uploads/2018/10/Groundswell-Saving-Lives-Saving-Money-Full-Report-Web-2016.pdf | UK (primary care + outpatient) |
| Fiori, K.P., Heller, C.G., Rehm, C.D., Parsons, A., Flattau, A., Braganza, S., Lue, K., Lauria, M., Racine, A. | Unmet Social Needs and No-Show Visits in Primary Care in a US Northeastern Urban Health System, 2018–2019 | 2020 | Am J Public Health. 2020;110:S242-S50 | USA (primary care) |
| Franciosi, E.B., Tan, A.J., Kassamali, B., Leonard, N., Zhou, G., Krueger, S., Rashighi, M., Lachance, A. | The Impact of Telehealth Implementation on Underserved Populations and No-Show Rates by Medical Specialty During the COVID-19 Pandemic | 2021 | Telemed J E Health. 2021 Aug 1;27(8):874-80 | USA (multiple) |
| Garuda, S. R., Javalgi, R.G., Talluri, V. S. | Tackling no-show behavior: a market-driven approach | 1998 | Health Mark Q. 1998;15(4):25-44 | n/a (review paper) |
| George, A., Rubin, G. | Non-attendance in general practice: a systematic review and its implications for access to primary health care | 2003 | Fam Pract. 2003;20(2):178-84 | n/a (review paper) |
| Godoy, L., Hodgkinson, S., Robertson, H.A., Sham, E., Druskin, L., Wambach, C.G., Beers, L.S. and Long, M. | Increasing Mental Health Engagement From Primary Care: The Potential Role of Family Navigation. | 2019 | Pediatrics. 2019;143(4):e20182418 | Review paper – primary care/mental health |
| Goldstein, E., Chokshi, B., Melendez-Torres, G.J., Rios, A., Jelley, M. and Lewis-O’Connor, A. | Effectiveness of trauma-informed care implementation in health care settings: Systematic review of reviews and realist synthesis. | 2024 | The Permanente Journal, 28(1), p.135. | Review paper – health general - trauma-informed practice |
| Gonzalez, J.S., Peyrot, M., McCarl, L.A., Collins, E.M., Serpa, L., Mimiaga, M.J., Safren, S.A. | Depression and Diabetes Treatment Nonadherence: A Meta-Analysis | 2008 | Diabetes Care. 2008;31(12):2398-403 | n/a (review paper) |
| Gray, S., Wells, K., Moodley, S., Rheuban, K. | Predictors of adolescent telemedicine visit no-shows during the covid-19 pandemic | 2022 | J Adolesc Health. 2022 Apr 1;70(4):S45. | USA (outpatient - adolescent health clinic) |
| Graham, E. | Staff Education Project Strategies for Reducing No-Shows | 2024 | Doctoral Dissertation, Walden University | USA – psychiatry |
| Gunner, E., Chandan, S.K., Marwick, S., Saunders, K., Burwood, S., Yahyouche, A., Paudyal, V | Provision and accessibility of primary healthcare services for people who are homeless: a qualitative study of patient perspectives in the UK | 2019 | Br J Gen Pract. 2019;69(685):e526-e36 | UK (primary care) |
| Guo, J.F., Bard, J.J., Morrice, D.R., Jaen, C., Poursani, R. | Offering transportation services to economically disadvantaged patients at a family health center: a case study | 2022 | Health Systems. 2022;11(4):251-75 | USA (primary care) |
| Gupta, A., Wagner, S., Raja, L., Struyven, R., Cortina-Borja, M., Keane, P. A., Huemer, J., Balaskas, K., Sim, D., Rahi, J., Solebo, A., Kang, S. | Determinants of Non-Attendance in Face-to-Face Ophthalmic Clinics Pre- and During the Coronavirus Pandemic | 2022 | 2022 Jun 1;63(7):2813-A0143. | UK (outpatient - ophthalmology) |
| Gurewich, D., Linsky, A.M., Harvey, K.L., Li, M., Griesemer, I., MacLaren, R.Z., Ostrow, R. and Mohr, D. | Relationship Between Unmet Social Needs and Care Access in a Veteran Cohort | 2023 | J Gen Intern Med. 2023;38(SUPPL 3):841-8 | USA (multiple - veterans administration) |
| Gurol-Urganci, I., de Jongh, T., Vodopivec-Jamsek, V., Atun, R., Car, J. | Mobile phone messaging reminders for attendance at healthcare appointments | 2013 | Cochrane Database of Systematic Reviews, 2013(12)(Art. No.: CD007458) | n/a (review paper) |
| Hamilton, W. | General practice non-attendance | 1999 | Br J Gen Pract 1999;49(445):664 | n/a (review paper) |
| Hamilton, W. | Non-attendance in general practice: a questionnaire | 2002 | Prim Health Care Res Dev. 2002;3(4):226-30 | UK (primary care) |
| Harrington, E.E., Reese‐Melancon, C. and Bock, J.E. | Sometimes they show, sometimes they don’t: Appointment attendance as a naturalistic prospective memory task | 2023 | Appl Cogn Psychol. 2023 May;37(3):590-9. | USA - college (non-medical study) |
| Haskell, T. and Cushman, M. | Process Improvement Initiatives to Reduce No Show Rates in Rural Communities. | 2020 | Circulation: Cardiovascular Quality and Outcomes, 13(Suppl_1)pp.A247-A247. | USA – paediatrics/rural health. |
| Healthwatch | Cost of living: People are increasingly avoiding NHS appointments and prescriptions | 2023 | https://www.healthwatch.co.uk/news/2023-01-09/cost-living-people-are-increasingly-avoiding-nhs-appointments-and-prescriptions | UK (health general) |
| Heidari, O., Winiker, A.K., Pollock, S., Sodder, S., Tsui, J.I. and Tobin, K.E. | A qualitative exploration of the use of telehealth for opioid treatment: Implications for nurse‐managed care. | 2024 | Journal of clinical nursing, 33(7), pp.2707-2718. | USA – substance use/telehealth |
| Henry, S. R., Goetz, M. B., Asch, S. M. | The Effect of Automated Telephone Appointment Reminders on HIV Primary Care No-Shows by Veterans | 2012 | J Assoc Nurses AIDS Care, 2012 Sep 1;23(5):409-18. | USA (primary care - HIV) |
| Herber, O.R., Jones, M.C., Smith, K., Johnston, D.W. | ‘Just not for me’ – contributing factors to nonattendance/ noncompletion at phase III cardiac rehabilitation in acute coronary syndrome patients: a qualitative enquiry | 2017 | J Clin Nurs. 2017;26(21-22):3529-42 | UK (outpatient - cardiac rehabilitation) |
| Hermoni, D., Mankuta, D., Reis, S. | Failure to Keep Appointments at a Community Health Centre: Analysis of causes | 1990 | Scandinavian journal of primary health care. 1990 Jan 1;8(2):107-11. | Israel (primary care) |
| Hickmott, S., Stroud, T. | Hidden Dimensions. The complexities of podiatry clinic non attendance of people with diabetes | 2009 | Diabet Med. 2009;26(SUPPL. 1):174. | UK (outpatient - diabetes) |
| Hilder, J., Gray, B. and Stubbe, M. | Health navigation and interpreting services for patients with limited English proficiency: a narrative literature review. | 2019 | Journal of primary health care, 11(3), pp.217-226. | Review paper – coordinators |
| Horigan, G., Davies, M., Findlay‐White, F., Chaney, D. and Coates, V. | Reasons why patients referred to diabetes education programmes choose not to attend: a systematic review | 2016 | Diabet Med. 2017;34(1):14-26. | n/a |
| Howarth, A. R., Apea, V., Michie, S., Morris, S., Sachikonye, M., Mercer, C. H., Evans, A., Delpech, V. C., Sabin, C., Burns, F. M. | Associations with sub-optimal clinic attendance and reasons for missed appointments among heterosexual women and men living with HIV in London | 2022 | AIDS Behav. 2022;26(11):3620-9. | UK (outpatient - HIV) |
| Howells, K., Amp, M., Burrows, M., Brown, J., Brennan, R., Dickinson, J., Jackson, S., Yeung, W.L., Ashcroft, D., Campbell, S. and Blakeman, T. | Remote primary care during the COVID-19 pandemic for people experiencing homelessness: a qualitative study. | 2022 | British Journal of General Practice, 72(720), pp.e492-e500. | UK (England) primary care/homelessness/telehealth |
| Huang, A. and Berg, W.T., | Telehealth: disparity and disappointment. | 2023 | Fertility and Sterility, 120(4), pp.817-818. | Opinion piece – telehealth |
| Hull, A. M., Alexander, D. A., Morrison, F., McKinnon, J. S. | A waste of time: non-attendance at out-patient clinics in a Scottish NHS Trust | 2002 | Health Bull (Edinb). 2002;60(1):62-9. | UK (outpatient - various) |
| Hussain-Gambles, M., Neal, R. D., Dempsey, O., Lawlor, D. A., Hodgson, J. | Missed appointments in primary care: questionnaire and focus group study of health professionals | 2004 | Br J Gen Pract. 2004;54(499):108-13. | UK (primary care) |
| Inglesfield, J. | Non-attendance and mental health problems in primary care | 1999 | Br J Gen Pract. 1999;49(443):488-9. | UK (primary care) |
| Izard, T. | Managing the Habitual No-Show Patient | 2005 | 2005 Feb;12(2):65-6. | USA (primary care) |
| Jackson, L.E. and Danila, M.I. | Healthcare disparities in telemedicine for rheumatology care. | 2022 | Current Opinion in Rheumatology, 34(3), pp.171-178 | USA – rheumatology/telehealth |
| Jefferson, L., Atkin, K., Sheridan, R., Oliver, S., Macleod, U., Hall, G., Forbes, S., Green, T., Allgar, V., Knapp, P. | Non-attendance at urgent referral appointments for suspected cancer: a qualitative study to gain understanding from patients and GPs | 2019 | Br J Gen Pract. 2019;69(689):E850-E9. | UK (outpatient - cancer referral) |
| Johnson, B. J., Mold, J. W.,Pontious, J. M. | Reduction and Management of No-Shows by Family Medicine Residency Practice Exemplars | 2007 | Annals Family Med. 2007;5(6):534-9 | USA (primary care) |
| Jones, M. C., Smith, K., Herber, O., White, M., Steele, F., & Johnston, D. W. | Intention, beliefs and mood assessed using electronic diaries predicts attendance at cardiac rehabilitation: An observational study | 2018 | Int J Nurs Stud. 2018;88:143-52. | UK (outpatient - cardiac rehabilitation) (Scotland) |
| Kannenberg, B. and Stadter, G. | Analysis and observations of telehealth in primary care follow up appointments for vulnerable populations. | 2022 | WMJ, 121(2), pp.116-120. | USA – primary care/telehealth |
| Kaplan-Lewis, E., Percac-Lima, S. | No-Show to Primary Care Appointments: Why Patients Do Not Come | 2013 | J Prim Care Community Health. 2013;4(4):251-5. | USA (primary care) |
| Kerr, G., Greenfield, G., Hayhoe, B., Gaughran, F., Halvorsrud, K., Pinto da Costa, M., Rehill, N., Raine, R., Majeed, A., Costelloe, C. and Neves, A.L. | Attendance at remote versus in-person outpatient appointments in an NHS Trust. | 2025 | Journal of Telemedicine and Telecare, 31(5), pp.721-731. | UK (England) – outpatient/remote care |
| Kielhold, K., Storholm, E.D., Reynolds, H.E., Vincent, W., Siconolfi, D.E., Kegeles, S.M., Pollack, L. and Campbell, C.K. | “I Don’t Feel Judged, I Don’t Feel Less of a Person”-Engaged and Supportive Providers in the HIV Care Experiences of Black Sexual Minority Men Living with HIV. | 2024 | Patient preference and adherence, pp.1641-1650. | USA – HIV care |
| Kiruparan, P., Kiruparan, N., Debnath, D. | Impact of pre-appointment contact and short message service alerts in reducing ‘Did Not Attend’ (DNA) rate on rapid access new patient breast clinics: a DGH perspective |  | BMC Health Serv Res. 2020;20(1):9. | UK (outpatient - new patient breast clinics) |
| Koester, K.A., Johnson, M.O., Wood, T., Fredericksen, R., Neilands, T.B., Sauceda, J., Crane, H.M., Mugavero, M.J., Christopoulos, K.A. | The influence of the ’good’ patient ideal on engagement in HIV care | 2019 | PLoS One. 2019;14(3). | USA (outpatient - HIV care) |
| Lacy, N. L., Paulman, A.,Reuter, M. D., Lovejoy, B. | Why We Don’t Come: Patient Perceptions on No-Shows | 2004 | Ann Fam Med. 2004;2(6):541-5. | USA (primary care) |
| Lakshminarayana, I. | Measures to improve non-attendance rates of community paediatric outpatient clinics | 2016 | Arch Dis Child. 2016;101(Supplement 1):A106 | UK (outpatient - community paediatrics) |
| Lasser, K. E., Mintzer, I. L., Lambert, A., Cabral, H., Bor, D. H. | Missed Appointment Rates in Primary Care: The Importance of Site of Care | 2005 | J Health Care Poor Underserved. 2005;16(3):475-86. | USA (primary care) |
| Lawal, M. O. | Non-attendance in diabetes education centres: perceptions of patients and education providers | 2014 | Diabet Med. 2014;31(SUPPL. 1):102-3. | UK (outpatient - diabetes education) |
| Lawal, M., & Woodman, A. | Socio-demographic Determinants of Attendance in Diabetes Education Centres: A Survey of Patients’ Views | 2021 | EMJ Diabetes. 2021;9(1):102-9. | UK (outpatient - diabetes education) |
| Leavey, G., Vallianatou, C., Johnson-Sabine, E., Rae, S., Gunputh, V. | Psychosocial Barriers to Engagement With an Eating Disorder Service: A Qualitative Analysis of Failure to Attend | 2011 | Eat Disord. 2011;19(5):425-40. | UK (outpatient - eating disorder) |
| Liu, S., Ng, J.K.Y., Moon, E.H., Morgan, D., Woodhouse, N., Agrawal, D., Chan, L., Chhabra, R. | Impact of COVID-19-associated anxiety on the adherence to intravitreal injection in patients with macular diseases a year after the initial outbreak | 2021 | Ther Adv Ophthalmol. 2022:1-12. | UK (outpatient - ophthalmology) |
| Logie, C.H. and Nyblade, L. | Recognizing and responding to stigma-related barriers in health care. | 2024 | Nature Reviews Disease Primers, 10(1), p.70. | Opinion/review paper – healthcare general |
| Lyon, R., Reeves, P. J. | An investigation into why patients do not attend for out-patient radiology appointments | 2005 | Radiography. 2006;12(4):283-90 | UK (outpatient - radiology) |
| Macharia, W.M. | An overview of interventions to improve compliance with appointment keeping for medical services | 1992 | JAMA. 1992;267(13):1813-7. | n/a (review paper) |
| Maehl, N., Bleckwenn, M., Riedel-Heller, S. G., Mehlhorn, S., Lippmann, S., Deutsch, T., Schrimpf, A. | The Impact of the COVID-19 Pandemic on Avoidance of Health Care, Symptom Severity, and Mental Well-Being in Patients With Coronary Artery Disease | 2021 | Front Med (Lausanne). 2021;8. | Germany (primary care) |
| Magan, T., Kirmani, A., Robertson, M., Mohamed, M., Mann, S. | Non-attendance in the ranibizumab treatment clinic for diabetic macular oedema: rates and reasons | 2014 | Eur J Ophthalmol. 2014;24(3):465-6. | UK (outpatient - ranibizumab treatment clinic) |
| Maggs, C., Langley, C. | Why patients miss primary care appointments: involving patients in research | 2008 | Prim Health Care. 2008;18(2):34-7. | UK (primary care) |
| Mahmood, F. | Exploring reasons for clients’ non-attendance at appointments within a community-based alcohol service: clients’ and practitioners’ perspectives. | 2021 | Doctoral dissertation, Manchester Metropolitan University | UK (outpatient - community-based alcohol services) |
| Margham, T. | Reducing missed appointments in general practice: evaluation of a quality improvement programme in East London. | 2021 | Br J Gen Pract. 2021;71(704):109- | UK (primary care) |
| Marshall, D., Quinn, C., Child, S., Shenton, D., Pooler, J., Forber, S., Byng, R. | What IAPT services can learn from those who do not attend | 2016 | J Ment Health. 2016;25(5):410-5. | UK (outpatient - IAPT) |
| Martin, C., Perfect, T., Mantle, G. | Non-attendance in primary care: the views of patients and practices on its causes, impact and solutions | 2005 | Fam Pract. 2005;22(6):638-43. | UK (primary care) |
| Martin, P.M. | Coroner inquest into 'hospital non-attendance' management in primary care | 2019 | Br J Gen Pract. 2019;69(681):195. | UK (primary care) |
| Martin, S. J., Bassi, S., Dunbar-Rees, R. | Commitments, norms and custard creams - a social influence approach to reducing did not attends (DNAs) | 2012 | J R Soc Med. 2012;105(3):101-4. | UK (primary care) |
| Mason, C. | Non-attendance at out-patient clinics: a case study | 1992 | J Adv Nurs. 1992;17(5):554-60. | UK (outpatient - various) |
| Masoud, T., Shah, A. and Joomun, S. | Reducing DNA Rates and Increasing Positive Contacts in an Outpatient Chronic Fatigue Service | 2017 | BMJ Quality Improvement Reports. 2017;6(1). | UK (outpatient - chronic fatigue service) |
| Maughan, D.L., Pearce, M. | Reducing non-attendance rates in community psychiatry: a case for sustainable development? | 2015 | BJPSych International, 2015;12(2):36–9. | UK (outpatient - community psychiatry) (England) |
| Mault, S., McDonough, B. J., Currie, P., Burhan, H. | Reasons proffered for non-attendance at a difficult asthma clinic | 2012 | Thorax. 2012;67(SUPPL. 2):A187. | UK (outpatient - asthma clinic) |
| Mayer, J., Abraham, P., Burhan, H., McDonough, B. J., Mault, S. | The effect of distance from the hospital, public transport availability and socioeconomic deprivation on non-attendance at a difficult asthma clinic | 2013 | Thorax. 2013;68:A198. | UK (outpatient - asthma clinic) |
| McCarthy, L., Parr, S., Green, S, Reeve, K. | Understanding models of support for people facing multiple disadvantage: A Literature Review | 2020 | https://www.shu.ac.uk/centre-regional-economic-social-research/publications/understanding-models-of-support-for-people-facing-multiple-disadvantage-a-literature-review | n/a (review paper) |
| McLean, S. M., Booth, A., Gee, M., Salway, S., Cobb, M., Bhanbhro, S., Nancarrow, S. A. | Appointment reminder systems are effective but not optimal: results of a systematic review and evidence synthesis employing realist principles | 2016 | Patient Prefer Adherence. 2016;10:479-99. | n/a (review paper) |
| McLean, S., Gee, M., Booth, A., Salway, S., Nancarrow, S., Cobb, M., Bhanbhro, S. | Targeting the use of reminders and notifications for uptake by populations (TURNUP): a systematic review and evidence synthesis | 2014 | Health Services and Delivery Research. 2014;2(34). | n/a (review paper) |
| McQueenie, R., Ellis, D. A., McConnachie, A., Wilson, P., Williamson, A. E. | Morbidity, mortality and missed appointments in healthcare: a national retrospective data linkage study | 2019 | BMC Med. 2019;17:9. | UK (primary care) |
| McQueenie, R., Ellis, D.A., Fleming, M., Wilson, P., Williamson, A.E. | Educational associations with missed GP appointments for patients under 35 years old: administrative data linkage study | 2021 | BMC Med. 2021 Sep 27;19(1):219 | UK (primary care) |
| Mehra, A., Hoogendoorn, C.J., Haggerty, G., Engelthaler, J., Gooden, S., Joseph, M., Carroll, S. and Guiney, P.A. | Reducing patient no-shows: an initiative at an integrated care teaching health center | 2018 | Journal of Osteopathic Medicine, 118(2), pp.77-84. | USA – osteopathic medicine |
| Milne, R.G | Reducing non-attendance at specialist clinics: an evaluation of the effectiveness and cost of patient-focussed booking and SMS reminders at a Scottish health board | 2010 | Int J Consum Stud. 2010;34(5):570-80. | UK (outpatient - various) |
| Minshall, I., Neligan, A. | A review of people who did not attend an epilepsy clinic and their clinical outcomes | 2017 | Seizure. 2017;50:121-4. | UK (primary care - epilepsy). |
| Mistry, S.K., Harris, E. and Harris, M. | Community health workers as healthcare navigators in primary care chronic disease management: a systematic review. | 2021 | Journal of general internal medicine, 36, pp.2755-2771. | Review paper – primary care/coordinators |
| Mistry, S.K., Shaw, M., Raffan, F., Johnson, G., Perren, K., Shoko, S., Harris-Roxas, B. and Haigh, F. | Inequity in access and delivery of virtual care interventions: a scoping review. | 2022 | International journal of environmental research and public health, 19(15), p.9411. | Review paper – virtual care |
| Mitchell, A.J., Selmes, Thomas | A Comparative Survey of Missed Initial and Follow-Up Appointments to Psychiatric Specialties in the United Kingdom | 2007 | Psychiatric services (Washington, DC). 2007;58(6):868-71. | UK (outpatient - psychiatry) |
| Morris, J., Campbell-Richards, D., Wherton, J., Sudra, R., Vijayaraghavan, S., Greenhalgh, T., Collard, A., Byrne, E., O'Shea, T. | Webcam consultations for diabetes: findings from four years of experience in Newham | 2017 | Pract Diabetes. 2017;34(2):45-50. | UK (outpatient - diabetes) |
| Morris, L., Haywood, S. | Why do patient miss appointments? A retrospective population study in paediatric outpatients in a metropolitan hospital | 2014 | Arch Dis Child. 2014;99(SUPPL. 1):A96. | UK (outpatient - paediatrics) |
| Moscrop, A. | Would it be a good idea to charge for missed appointments at the doctors surgery? | 2015 | BMJ Opinion. 2015;351:23-. | n/a (opinion) |
| Moscrop, A., Siskind, D., Stevens, R. | Mental health of young adult patients who do not attend appointments in primary care: a retrospective cohort study | 2012 | Fam Pract. 2012;29(1):24-9. | UK (primary care) |
| Murray, M. | Modernising the NHS - Patient care: access | 2000 | BMJ 2000 Jun 10;320(7249):1594-6 | UK (health general) |
| Nancarrow, S., Bradbury, J., Avila, C. | Factors associated with non-attendance in a general practice super clinic population in regional Australia: A retrospective cohort study | 2014 | Australas Med J. 2014;7(8):323-33 | Australia (primary care) |
| Neal, R. D., Lawlor, D. A., Allgar, V., Colledge, M., Ali, S., Hassey, A., Portz, C., Wilson, A. | Missed appointments in general practice: retrospective data analysis from four practices | 2001 | Br J Gen Pract. 2001;51(471):830-2 | UK (primary care) |
| Neal, R.D., Hussain-Gambles, M., Allgar, V.L., Lawlor, D.A., Dempsey, O. | Reasons for and consequences of missed appointments in general practice in the UK: questionnaire survey and prospective review of medical records | 2005 | BMC Fam Pract. 2005;6:47 | UK (primary care) |
| Nguyen, D.L., Dejesus, R.S., Wieland, ML. | Missed Appointments in Resident Continuity Clinic: Patient Characteristics and Health Care Outcomes | 2011 | J Grad Med Educ. 2011;3(3):350-5 | USA (primary care) |
| NHS England | Approaches to implementing two-way appointment reminders | 2023 | NHS England | UK (health general) |
| Ogunyemi, A.O. | Reducing the Prevalence of Missed Primary Care Appointments in Community Health Centers. Capstone Project Paper | 2020 | Doctoral Dissertation, University of Southern California | USA (primary care) |
| Opon, S., Ochieng, T., Wanja M., Njoroge, K.M. | The effect of patient reminders in reducing missed appointment in medical settings: a systematic review | 2020 | PAMJ-One Health, 2(9) | n/a (review paper) |
| Pakhomova, T.E., Nicholson, V., Fischer, M., Ferguson, J., Moore, D.M., Salters, K., Lester, R.T., Kremer, H., Dawydiuk, N., Barrios, R. and Parashar, S. | Exploring Primary Healthcare Experiences and Interest in Mobile Technology Engagement Amongst an Urban Population Experiencing Barriers to Care | 2023 | Qual Health Res. 2023;33(8-9):765-77 | Canada (primary care) |
| Pal, B., Taberner, D. A., Readman, L. P., Jones, P. | Why do outpatients fail to keep their clinic appointments? Results from a survey and recommended remedial actions | 1998 | Int J Clin Pract. 1998;52(6):436-7. | UK (outpatient - various) |
| Parker, M.M., Moffet, H.H., Schillinger, D., Adler, N., Fernandez, A., Ciechanowski, P., Karter, A.J. | Ethnic Differences in Appointment Keeping and Implications for the Patient-Centered Medical Home - Findings from the Diabetes Study of Northern California (DISTANCE) | 2012 | Health Serv Res. 2012;47(2):572-93. | USA (primary care - diabetes) |
| Parkes, T., Matheson, C., Carver, H., Foster, R., Budd, J., Liddell, D., Wallace, J., Pauly, B., Fotopoulou, M., Burley, A., Anderson, I., MacLennan, G. | A peer-delivered intervention to reduce harm and improve the well-being of homeless people with problem substance use: the SHARPS feasibility mixed-methods study | 2022 | Health Technol Assess. 2022;26(14):1-128. | UK (outreach care) |
| Parsons, J., Abel, G., Mounce, L.T., Atherton, H. | The changing face of missed appointments | 2023 | Br J Gen Pract. 2023;73(728):134-5. | UK (general) |
| Parsons, J., Bryce, C., Atherton, H. | Which patients miss appointments with general practice and the reasons why: a systematic review | 2021 | Br J Gen Pract. 2021;71(707):E406-E12. | n/a (review paper) |
| Peart A., Lewis V., Brown T., and Russell, G. | Patient navigators facilitating access to primary care: a scoping review. | 2018 | BMJ Open 2018;8:e019252. | Review paper – primary care/coordinators |
| Perron, N.J., Dao, M.D., Kossovsky, M.P., Miserez, V., Chuard, C., Calmy, A., Gaspoz, J-M. | Reduction of missed appointments at an urban primary care clinic: a randomised controlled study | 2010 | BMC Fam Pract. 2010;11. | Switzerland (primary care) |
| Poll, R., Allmark, P., Tod, A. M. | Reasons for missed appointments with a hepatitis C outreach clinic: A qualitative study | 2017 | Int J Drug Policy. 2017;39:130-7. | UK (outpatient - hepatitis C) |
| Potter, L.C., Stone, T., Swede, J., Connell, F., Cramer, H., McGeown, H., Carvalho, M., Horwood, J., Feder, G. and Farr, M. | Improving access to general practice for and with people with severe and multiple disadvantage: a qualitative study. | 2024 | British Journal of General Practice 74 (742): e330-e338 | UK (England) – primary care |
| Practice Nurse (no author named) | Missing appointments increases risk of death | 2019 | Practice Nurse. 2019:49(1) | UK (primary care) |
| Practice Nurse (no author named) | Change booking system to cut DNAs | 2020 | Practice Nurse. 2020:50(10) | UK (primary care) |
| Prentice, P | Missed appointments | 2004 | Practice Management.2004:14(8) | UK (primary care) |
| Prudden, G. | Quality improvement project exploring the factors in non-attendance at an NHS musculoskeletal outpatients department | 2021 | Physiotherapy (United Kingdom). 2021;113(Supplement 1):e151-e2. | UK (outpatient - musculoskeletal) |
| Qin, J., Chan, C.W., Dong, J., Homma, S. and Ye, S. | Telemedicine is associated with reduced socioeconomic disparities in outpatient clinic no-show rates | 2023 | Journal of Telemedicine and Telecare. 2023:0(0) | USA (outpatient - internal medicine) |
| Raja, L., Wagner, S., Struyven, R., Cortina-Borja, M., Keane, P. A., Huemer, J., Balaskas, K., Sim, D., Rahi, J., Solebo, A. L., Kang, S. | Determinants of non-attendance in synchronous teleophthalmology clinics. | 2022 | Investigative Ophthalmology and Visual Science. 2022:63(7):1412-A0108. | UK (outpatient - ophthalmology) |
| Reekie, D., Devlin, H. | Preventing failed appointments in general dental practice: a comparison of reminder methods | 1998 | Br Dent J. 1998;185(9):472-4. | UK (primary care - dental) |
| Revolving Doors Agency | Navigating complexity: learning from Navigators across Birmingham | 2020 | https://www.tnlcommunityfund.org.uk/media/insights/documents/Navigating-Complexity-Learning-from-the-navigators-across-Birmingham-2020.pdf?mtime=20220601114929&focal=none | UK (non-health setting) |
| Roberts, L., Garo-Falides, J., Bowran, H. | Non-attendance in musculoskeletal outpatients: The good, the bad and the ugly | 2015 | Physiotherapy (United Kingdom). 2015;101(SUPPL. 1):eS1289-eS90. | UK (outpatient - musculoskeletal) |
| Robotham, D., Satkunanathan, S., Reynolds, J., Stahl, D., Wykes, T. | Using digital notifications to improve attendance in clinic: systematic review and meta-analysis | 2016 | BMJ Open. 2016;6(10):14. | n/a (review paper) |
| Rose, K.D., Ross, J.S., Horwitz, LI.. | Advanced access scheduling outcomes: a systematic review | 2011 | Archives of Internal Medicine, 2022:171(13) | n/a (review paper) |
| Ross, S. K. | Cancellation and default from appointments in primary care | 1991 | Br J Gen Pract. 1991;41(342):34. | UK (primary care) |
| Rowett, M., Reda, S., Makhoul, S. | Prompts to Encourage Appointment Attendance for People With Serious Mental Illness | 2010 | Schizophr Bull. 2010;36(5):910-1. | n/a (review paper) |
| Royal College of General  Practitioners | Missed GP appointments are frustrating – but there may be underlying reasons why patients don't turn up, says College | 2020 | https://www.rcgp.org.uk/news/missed-gp-appointments | UK (primary care) |
| Royal College of General Practitioners | Charging for missed appointments won’t address intense GP pressures | 2022 | https://www.rcgp.org.uk/news/missed-appointments | UK (primary care) |
| Royal College of General Practitioners | Charging for GP appointments would have the biggest impact on vulnerable patients, says College Chair | 2023 | https://www.rcgp.org.uk/News/GP-appointment-charges-response | UK (primary care) |
| Ruggeri, K., Folke, T., Benzerga, A., Verra, S., Buttner, C., Steinbeck, V., Yee, S., Chaiyachati, K. | Nudging New York: adaptive models and the limits of behavioral interventions to reduce no-shows and health inequalities | 2020 | BMC Health Serv Res. 2020;20(1). | USA (multiple) |
| Samuels, R.C., Ward, V.L., Melvin, P., Macht-Greenberg, M., Wenren, L. M., Yi, J., Massey, G., Cox, J.E. | Missed Appointments: Factors Contributing to High No-Show Rates in an Urban Pediatrics Primary Care Clinic | 2015 | Clin Pediatr. 2015;54(10):976-82. | USA (primary care - peadiatric) |
| Samyn, M., Fihosy, S., Day, J.M. and Hames, A., | Showing we care: reducing non-attendance rates in an adolescent clinic. | 2018 | Arch Dis Child 2018;0:1. | UK (England) – adolescent clinic |
| Schwebel, F.J., Larimer, M. E. | Using text message reminders in health care services: A narrative literature review | 2018 | Internet Interv. 2018;13:82-104. | n/a (review paper) |
| Scottish Government | Inclusion Health Action in General Practice: Early Evaluation Report | 2024 | https://www.gov.scot/publications/inclusion-health-action-general-practice-early-evaluation-report/ | UK (Scotland) – primary care |
| Shah, S.J., Cronin, P., Hong, C.S., Hwang, A.S., Ashburner, J.M., Bearnot, B.I., Richardson, C.A., Fosburgh, B.W. and Kimball, A.B. | Targeted Reminder Phone Calls to Patients at High Risk of No-Show for Primary Care Appointment: A Randomized Trial | 2016 | J Gen Intern Med. 2016;31(12):1460-6 | USA (primary care) |
| Shahab, I., Meili, R. | Examining non-attendance of doctor’s appointments at a community clinic in Saskatoon | 2019 | Can Fam Physician. 2019;65(6):E264-E8 | Canada (primary care) |
| Shao, C.C., Katta, M.H., Smith, B.P., Jones, B.A., Gleason, L.T., Abbas, A., Wadhwani, N., Wallace, E.L., Mugavero, M.J. and Chu, D.I. | Reducing no-show visits and disparities in access: the impact of telemedicine. | 2024 | Journal of Telemedicine and Telecare, 2024;0(0). | USA – tertiary academic center/telehealth |
| Sharp, D. J., Hamilton, W. | Non-attendance at general practices and outpatient clinics | 2001 | BMJ (Clinical research ed). 2001;323(7321):1081-2. | UK (primary care + outpatient) |
| Sharp, L., Cotton, S., Thornton, A., Gray, N., Cruickshank, M., Whynes, D., Duncan, I., Hammond, R., Smart, L., Little, J., Tombola Grp | Who defaults from colposcopy? A multi-centre, population-based, prospective cohort study of predictors of non-attendance for follow-up among women with low-grade abnormal cervical cytology | 2012 | European Journal of Obstetrics & Gynecology and Reproductive Biology. 2012;165(2):318-25. | UK (outpatient - colposcopy) |
| Sheffield Children’s NHS Foundation Trust/NHS Strategy Unit | Was Not Brought, Sheffield Children’s NHS Foundation Trust | 2024 | https://www.strategyunitwm.nhs.uk/sites/default/files/2023-04/Was%20Not%20Brought%2C%20Sheffield%20Children%E2%80%99s%20NHS%20Foundation%20Trust%20_V1.0_FINAL.pdf | UK (England) – paediatric care |
| Shimotsu, S., Roehrl, A., McCarty, M., Vickery, K., Guzman-Corrales, L.,Linzer, M., Garrett, N. | Increased Likelihood of Missed Appointments (“No Shows”) for Racial/Ethnic Minorities in a Safety Net Health System | 2016 | J Prim Care Community Health. 2016;7(1):38-40 | USA (multiple) |
| Shommu, N.S., Ahmed, S., Rumana, N., Barron, G.R., McBrien, K.A. and Turin, T.C. | What is the scope of improving immigrant and ethnic minority healthcare using community navigators: a systematic scoping review. | 2016 | International journal for equity in health, 15, pp.1-12. | Review paper – immigrant/minority healthcare/coordinators. |
| Simmons, D., Clover, G. | A case control study of diabetic patients who default from primary care in urban New Zealand | 2007 | Diabetes Metab. 2007;33(2):109-13. | New Zealand (primary care - diabetes) |
| Sims, H., Sanghara, H., Hayes, D., Wandiembe, S., Finch, M., Jakobsen, H., Tsakanikos, E., Okocha, C.I. and Kravariti, E., | Text Message Reminders of Appointments: A Pilot Intervention at Four Community Mental Health Clinics in London | 2012 | Psychiatr Serv. 2012 Feb;63(2):161-8 | UK (outpatient - community mental health) |
| Smith, E. and Badowski, M.E. | Telemedicine for HIV care: current status and future prospects. | 2021 | HIV/AIDS-Research and Palliative Care, pp.651-656. | Review article – HIV care |
| Smith, L.B., Yang, Z., Golberstein, E., Huckfeldt, P., Mehrotra, A. and Neprash, H.T. | The effect of a public transportation expansion on no‐show appointments. | 2022 | Health services research, 57(3), pp.472-481. | USA – healthcare general |
| Stevenson, J. S. | Appointment systems in general practice: How patients use them | 1967 | BMJ 1967 Jun 6;2(5555):827. | UK (primary care) |
| Sumarsono, A., Case, M., Kassa, S. and Moran, B., | Telehealth as a Tool to Improve Access and Reduce No‑Show Rates in a Large Safety‑Net Population in the USA | 2023 | Bull N Y Acad Med. 2023;100(2):398-407 | USA (various) |
| Sun, C.A., Shenk, Z., Renda, S., Maruthur, N., Zheng, S., Perrin, N., Levin, S. and Han, H.R. | Experiences and Perceptions of Telehealth Visits in Diabetes Care During and After the COVID-19 Pandemic Among Adults With Type 2 Diabetes and Their Providers: Qualitative Study | 2023 | JMIR diabetes. 2023;8:e44283-e | USA (outpatient - diabetes care).` |
| Sun, C-A, Taylor, K., Levin, S., Renda, S.M., Han, H-R. | Factors associated with missed appointments by adults with type 2 diabetes mellitus: a systematic review | 2021 | BMJ Open Diabetes Res Care. 2021;9(1) | n/a (review paper) |
| Tait, J., Noyes, K., Bath, L., Henderson, M., Elleri, D. | Clinic non-attendance, glycaemic control and deprivation score in paediatric and young persons' diabetes clinics in Lothian, Scotland | 2017 | Pediatr Diabetes. 2017;18(Supplement 25):88. | UK (outpatient - paediatric/young persons diabetes clinic) |
| Taylor, B. | Patient use of a mixed appointment system in an urban practice | 1984 | BMJ. 1984;289(6454):1277-8. | UK (primary care) |
| Teggart, K., Neil-Sztramko, S.E., Nadarajah, A., Wang, A., Moore, C., Carter, N., Adams, J., Jain, K., Petrie, P., Alshaikhahmed, A. and Yugendranag, S. | Effectiveness of system navigation programs linking primary care with community-based health and social services: a systematic review. | 2023 | BMC Health Services Research, 23(1), p.450. | Review paper – community-based health and social services/coordinators. |
| Tello, Y. and Gianelis, K.A. | Improving Equitable Postpartum Care in an Urban Private Clinic with Predominantly Black Patients. | 2024 | Journal of Midwifery & Women's Health, 69(5), pp.784-789. | USA – postpartum care |
| Teo, A.R., Niederhausen, M., Handley, R., Metcalf, E.E., Call, A.A., Jacob, R.L., Zikmund-Fisher, B.J., Dobscha, S.K. and Kaboli, P.J. | Using Nudges to Reduce Missed Appointments in Primary Care and Mental Health: a Pragmatic Trial | 2023 | Journal of General Internal Medicine. 2023;38(SUPPL 3):894-904. | USA (primary care) |
| Thapar, A,. Ghosh, A. | Non-attendance at a psychiatric clinic | 1991 | Psychiatr Bull. 1991;15(4):205-6. | UK (outpatient - psychiatry) |
| Tierney, A.A., Mosqueda, M., Cesena, G., Frehn, J.L., Payan, D.D. and Rodriguez, H.P. | Telemedicine implementation for safety net populations: a systematic review. | 2024 | Telemedicine and e-Health, 30(3), pp.622-641. | Review paper - telehealth |
| Tonnesen, M., Hedeager Momsen, A.M. | Bridging gaps in health? A qualitative study about bridge-building and social inequity in Danish healthcare | 2023 | Int J Qual Stud Health Well-being 2023;18(1):2241235. | Denmark (health general) |
| Traeger, L., O'Cleirigh, C.l., Skeer, M.R., Mayer, K.H., Safren, S.A. | Risk factors for missed HIV primary care visits among men who have sex with men | 2012 | J Behav Med. 2012;35(5):548-56 | USA (primary care - HIV) |
| Ullah, S., Rajan, S., Liu, T., Demagistris, E., Jahrstorfer, R., Anandan, S., Gentile, C., Gill, A. | Why do Patients Miss their Appointments at Primary Care Clinics? | 2018 | J Fam Med Dis Prev. 2018;4(3):1-5. | USA (primary care) |
| Unger, K., Lesiuk, A. and Unger, S. | How do we save £1200 lost to dnas’ per complex paediatric respiratory clinic and protect our most vulnerable patients? | 2023 | Arch Dis Child 2023;108:A441 | UK (outpatient - complex paediatric respiratory clinic) |
| NHS | Identifying causes of patient Did Not Attends (DNAs) | 2023 | NHS England | UK (health general) |
| NHS | Identifying causes of patient Did Not Attends (DNAs) – example script | 2023 | NHS England | UK (health general) |
| Valaitis, R.K., Carter, N., Lam, A., Nicholl, J., Feather, J. and Cleghorn, L., 2017. | Implementation and maintenance of patient navigation programs linking primary care with community-based health and social services: a scoping literature review. | 2017 | BMC health services research, 17, pp.1-14. | Review paper – community-based health and social services/coordinators |
| van Baar, J. D.,Joosten, H.,Car, J.,Freeman, G. K.,Partridge, M. R.,van Weel, C.,Sheikh, A. | Understanding reasons for asthma outpatient (non)attendance and exploring the role of telephone and e-consulting in facilitating access to care: exploratory qualitative study | 2006 | Qual Saf Health Care. 2006;15(3):191-5. | UK (outpatient - asthma) |
| Verity, A. and Brown, V.T. | GP access for inclusion health groups: perspectives and recommendations. | 2024 | BJGP open, 8(3). | UK (England) primary care |
| Vetter, I. | Primary health care for people with multiple and complex needs: what does best practice look like? | 2020 | https://www.bht.org.uk/wp-content/uploads/2021/02/Primary-Healthcare-What-does-best-practice-look-like-May-2020.pdf | UK (primary care) |
| Waid, J., Halpin, K. and Donaldson, R. | Mental health service navigation: a scoping review of programmatic features and research evidence. | 2021 | Social Work in Mental Health, 19(1), pp.60-79. | Review paper – mental health/coordination |
| Waller, J., Hodgkin, P. | Defaulters in general practice: who are they and what can be done about them? | 2000 | Fam Pract. 2000;17(3):252-3 | UK (primary care) |
| Wang, Y., Baidoo, F.A. | Design of Integral Reminder for Collaborative Appointment Management | 2017 | 50^th^ Annual Hawaii International Conference on System Sciences (HICSS). 2017:910-9. | n/a (other) |
| Wells, J.P. and Sivarajasingam, V. | Improving attendance: an audit of missed appointments with two successful interventions. | 2011 | British Journal of Oral and Maxillofacial Surgery, 49, p.S114. | UK (Wales) - oral and maxillofacial surgery |
| Weltermann, B.M., Doost, S.M., Kersting, C., Gesenhues, S. | Hypertension management in primary care: how effective is a telephone recall for patients with low appointment adherence in a practice setting? | 2014 | Wiener klinische Wien Klin Wochenschr. 2014;126(19-20):613-8. | Germany (primary care - hypertension) |
| Wilkinson, M. J. | Effecting change in frequent non-attenders | 1994 | Br J Gen Pract. 1994 May;44(382):233 | UK (primary care) |
| Williamson, A. E., Ellis, D.A., Wilson, P., McQueenie, R., McConnachie, A. | Understanding repeated non-attendance in health services: a pilot analysis of administrative data and full study protocol for a national retrospective cohort | 2017 | BMJ Open. 2017;7(2):11 | UK (primary care) |
| Williamson, A.E., McQueenie, R., Ellis, D.A., McConnachie, A., Wilson, P. | Missingness' in health care: Associations between hospital utilization and missed appointments in general practice. A retrospective cohort study | 2021 | PLoS One. 2021;16(6):e0253163. | UK (primary care) |
| Wilsey, K.L. | Why Patients Miss Appointments at an Integrated Primary Care Clinic | 2020 | Doctoral Dissertation, Antioch University | USA (primary care) |
| Wilson, B., Astley, P. | Gatekeepers: Access to Primary Care for those with Multiple Needs | 2016 | Stoke-on-Trent: VOICES, Healthwatch and Expert Citizens CIC | UK (primary care) |
| Wilson, J., Lau, D., Kristoferson, E., Ginzler, E. and Kabani, N. | A patient-centered evaluation of a novel medical student-based patient navigation program. | 2024 | Patient education and counseling, 120, p.108131. | USA – rheumatology/coordinators |
| Wilson, R., Winnard, Y. | Causes, impacts and possible mitigation of non-attendance of appointments within the National Health Service: a literature review | 2022 | J Health Organ Manag. 2022;36(7):892-911. | UK (health general) |
| Winkley, K., Evwierhoma, C., Amiel, S. A., Lempp, H. K., Ismail, K., Forbes, A. | Patient explanations for non-attendance at structured diabetes education sessions for newly diagnosed Type 2 diabetes: a qualitative study | 2015 | Diabet Med. 2015;32(1):120-8. | UK (outpatient - diabetes education) |
| Woodcock, E.W. | Managing your appointment 'no-shows' | 2000 | J Med Pract Manag. 2000;15:284-8. | USA (primary care) |
| Yates, L., Brittleton, L., Bean, N. | An investigation into the factors which inﬂuence attendance rates for psychology appointments in an adult intellectual disability service | 2022 | Adv Ment Health Intellect Disabil. 2022 Aug 31;16(4):216-25 | UK (outpatient - psychology/intellectual disability) |
| Zailinawati, A. H., Ng, C. J., Nik-Sherina, H. | Why do patients with chronic illnesses fail to keep their appointments? A telephone interview | 2006 | Asia-Pac J Public Health. 2006;18(1):10-5. | Malaysia (primary care) |
| Zulman, D.M., O’Brien, C.W., Slightam, C., Breland, J.Y., Krauth, D. and Nevedal, A.L. | Engaging high-need patients in intensive outpatient programs: a qualitative synthesis of engagement strategies. | 2018 | Journal of General Internal Medicine, 33, pp.1937-1944. | USA – outpatient/secondary care. |

### 5. GUIDED checklist (76)

| **Item description** | **Page in manuscript where item is located** | **Other locations** |
| --- | --- | --- |
| 1. Report the context for which the intervention was developed. | pp.4-5 | Lindsay et al, 2023  Lindsay et al, 2024 |
| 2. Report the purpose of the intervention development process. | pp.4-5 | Lindsay et al, 2024 |
| 3. Report the target population for the intervention development process. | p.3 | Lindsay et al, 2024 |
| 4. Report how any published intervention development approach contributed to the development process. | p.5 |  |
| 5. Report how evidence from different sources informed the intervention development process. | Pp6-8  Table 1. |  |
| 6. Report how/if published theory informed the intervention development process. | p.9 | Baruffati et al **forthcoming.** |
| 7. Report any use of components from an existing intervention in the current intervention development process. | Supporting information, section 4. |  |
| 8. Report any guiding principles, people or factors that were prioritised when making decisions during the intervention development process. | p.10  Figure 1.  Supporting information, section 6. |  |
| Report how stakeholders contributed to the intervention development process. | pp.5-8; Table 1  Supporting information, section 6. |  |
| 10. Report how the intervention changed in content and format from the start of the intervention development process. | p.9  Supporting information, section 6. |  |
| 11. Report any changes to interventions required or likely to be required for subgroups. | p.16 |  |
| 12. Report important uncertainties at the end of the intervention development process. | pp.27-28 |  |
| 13. Follow TIDieR guidance when describing the developed intervention. | n/a | For future reporting. |
| 14. Report the intervention development process in an open access format. | Supporting information, section 6. | OSF page  Final report (forthcoming) |

### 6. Stakeholder Advisory Group workshop materials

#### 6.1 Workshop 1 (online)

**Aims**:

- To refine and validate our final programme theory on the causes of missingness.
- To outline initial actions to address these causes at different ‘levels’ of the social-ecological model.

After a presentation of the main findings of our review into the causes of missingness, the stakeholder group was divided into four subgroups to discuss fictional case studies, an example of which is below in figure 1. Case studies were sent to participants prior to the meeting. Groups were asked two questions:

1. What parts of our programme theory are in this person’s story?
2. Does the programme theory represent a clear, accurate and adequate description of missingness? Who or what is missing?

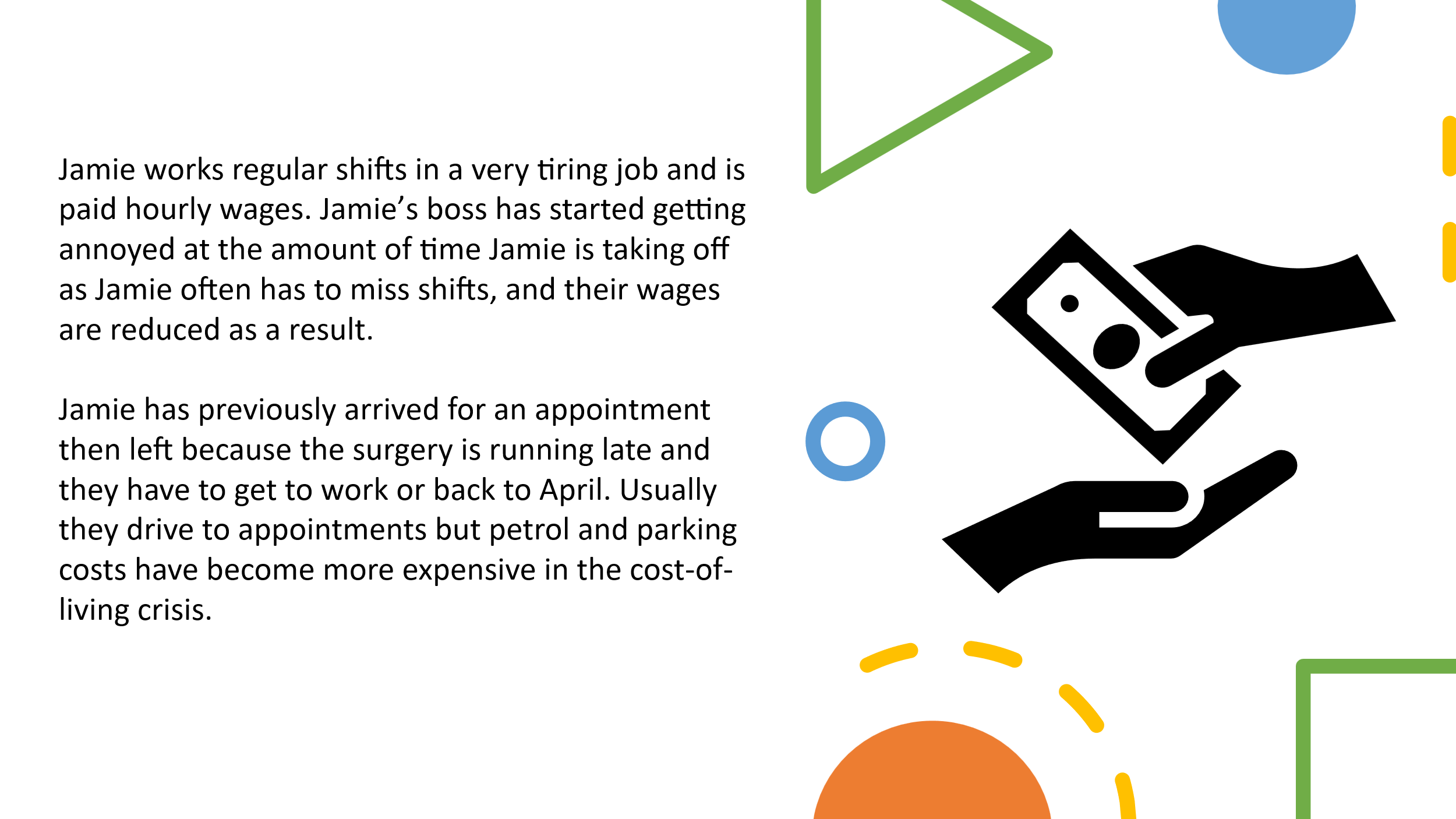

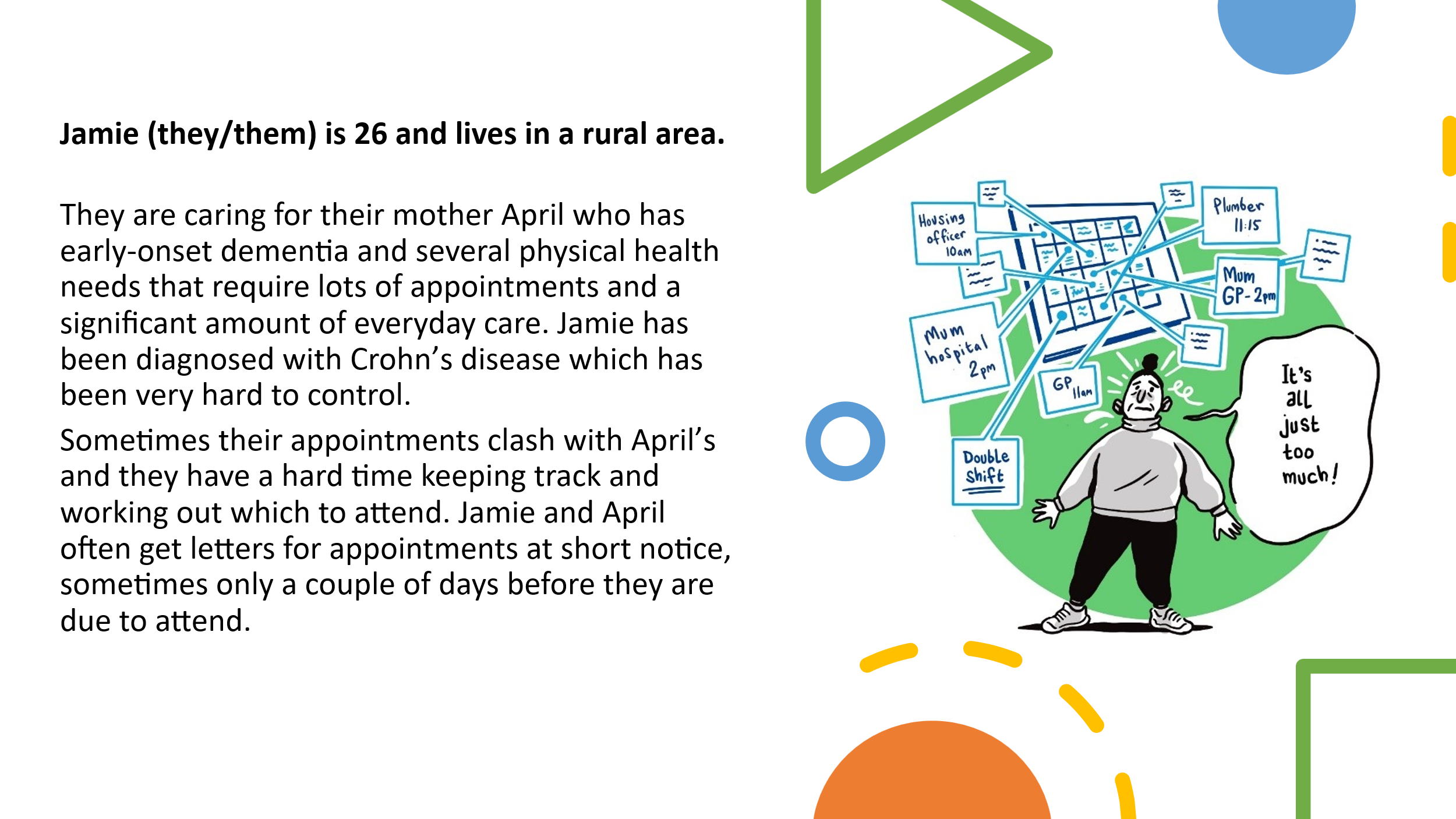

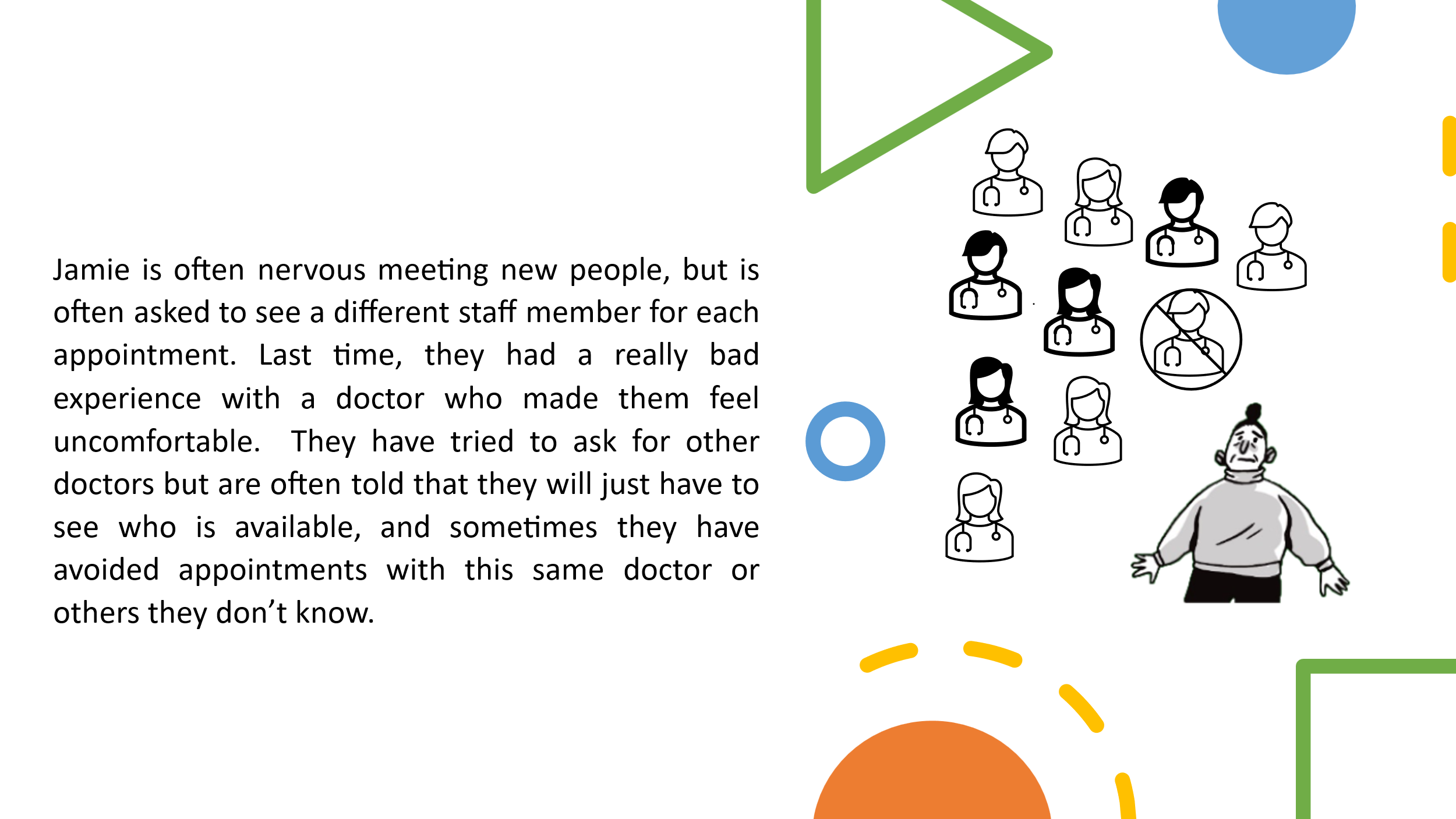

Figure 1: Example case study

In a second activity, groups were asked to identify possible interventions or actions that might make it easier for people to attend appointments. These were matched with areas of the programme theory, then places on a diagram of the social-ecological model to show which ‘level’ they would involve. The board and a set of notes are in figure 2.

Figure 2: Template for online discussion using the social-ecological model

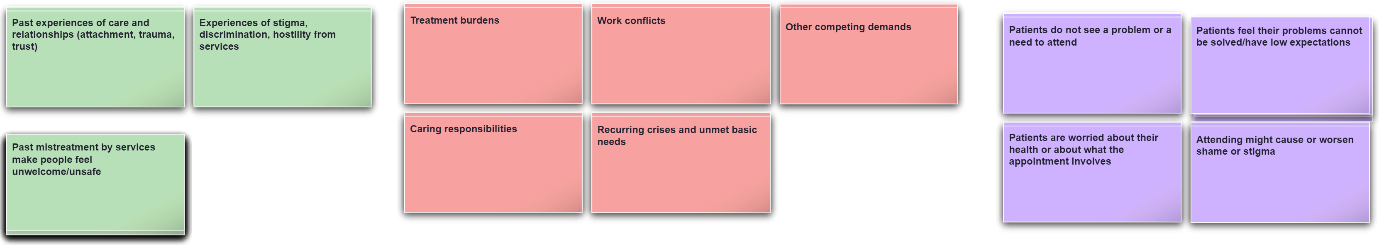

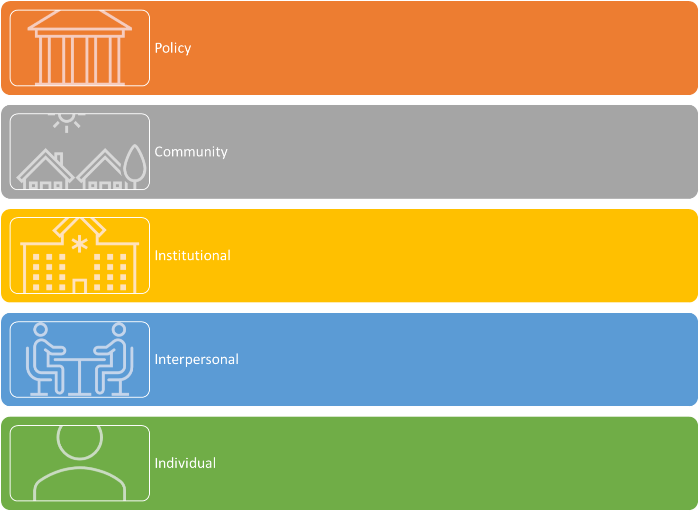

#### 6.2 Workshop 2 (in-person)

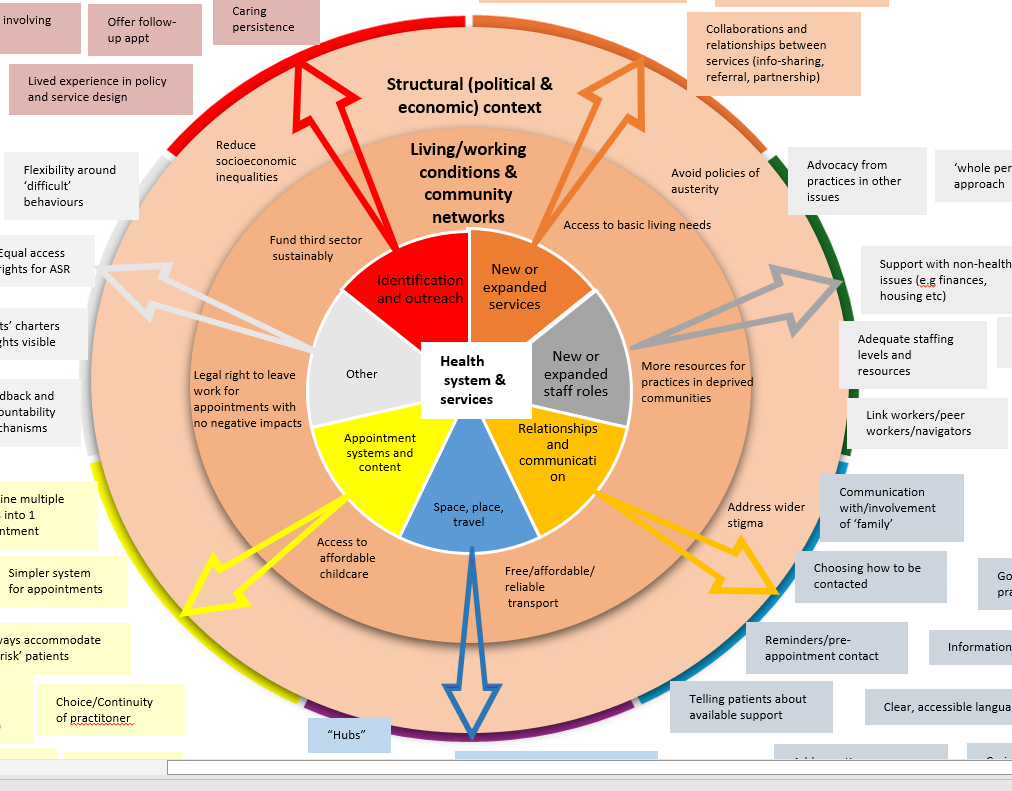
The research team collated intervention ideas from the first StAG with suggestions from the realist review and the interviews, then outlined a set of wider domains: appointment systems and content; space, place and travel; relationships and communication; new or expanded staff roles; new or expanded services; identification and outreach; other. These were placed on a diagram again showing contextual ‘levels’, with those activities already suggested placed on the diagram. The template is in figure 3. Participants were asked the following questions:

Figure 3: Template for discussion of intervention activities at different 'levels'

- What actions could we take in specific domains to address missingness?
- Are there specific interventions that would help with some of the causes identified in our theory?

In the second activity, the StAG members were asked to outline and then rank the core principles underpinning any intervention activity, both adding their own contributions and critiquing a set of principles already identified in the other work packages. Through some refinement by the research team, these principles became the principles of the missingness lens outlined in the paper.

#### 6.3 Workshop 3 (in-person)

Reviewing the findings of the workshops to date, and the findings of ongoing interviews and literature review, the research team refined the intervention domains to the final domains outlined here. The goal was then to refine specific actions or approaches within these domains, linked to the causes of missingness, and to explore possible drawbacks, challenges or obstacles to success. We did this through Round Robin Brainstorming. Participants were divided into small groups and given responsibility for designing actions within an intervention domain. Another group would then critique this idea, and the first group would respond by refining their intervention further. After two rounds of critique and refined, each domain would have a final set of actions alongside critiques, key consideration, essential resources and possible drawbacks or unintended outcomes. The template for this process is in figure 4.

Figure 4: example template for refined activities in the domain of relationships and communication

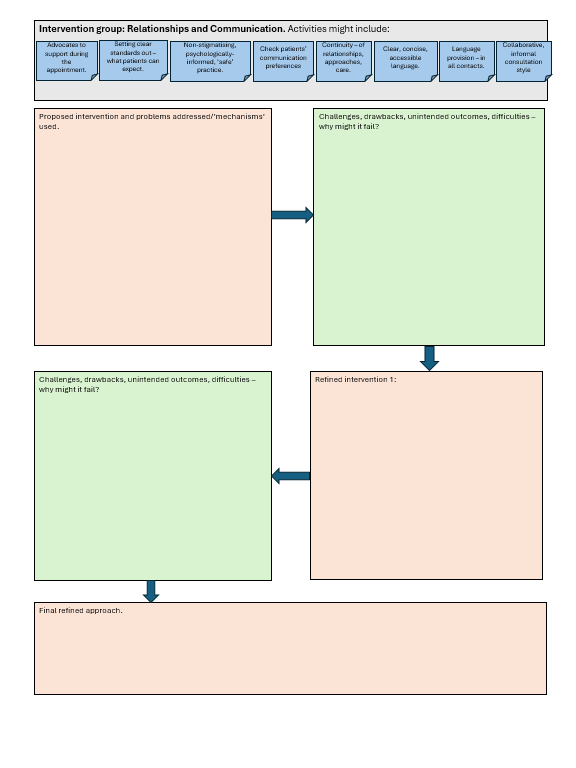

#### 6.4 Workshop 4 (in-person)

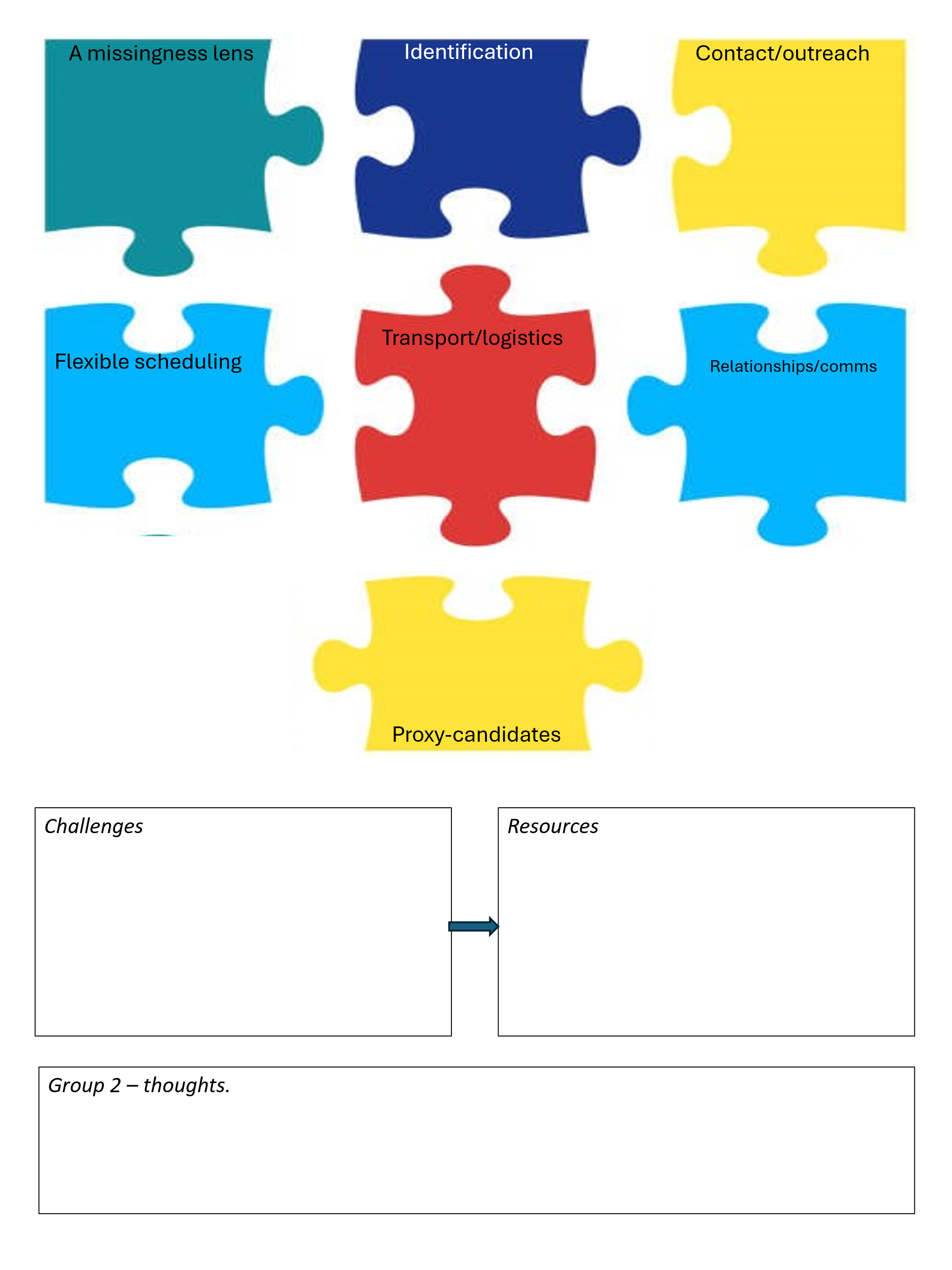
The final workshop was designed to finalise the intervention and any further considerations for its implementation. Again using small groups, we utilised fictional case studies of services (one Deep End practice, one ‘mainstream’ practice and one inclusion health practice) and asked small groups to apply the different domains of the intervention to them. Groups would then discuss the challenges the practice might face implementing their interventions, and how they might overcome this challenge either by changing their approach or by applying additional resources. A second group would then review the intervention and discuss whether they felt it would work, whether it was inclusive to people experiencing missingness, and whether it would be used. An example case study and template are included in figure 5 and 6.

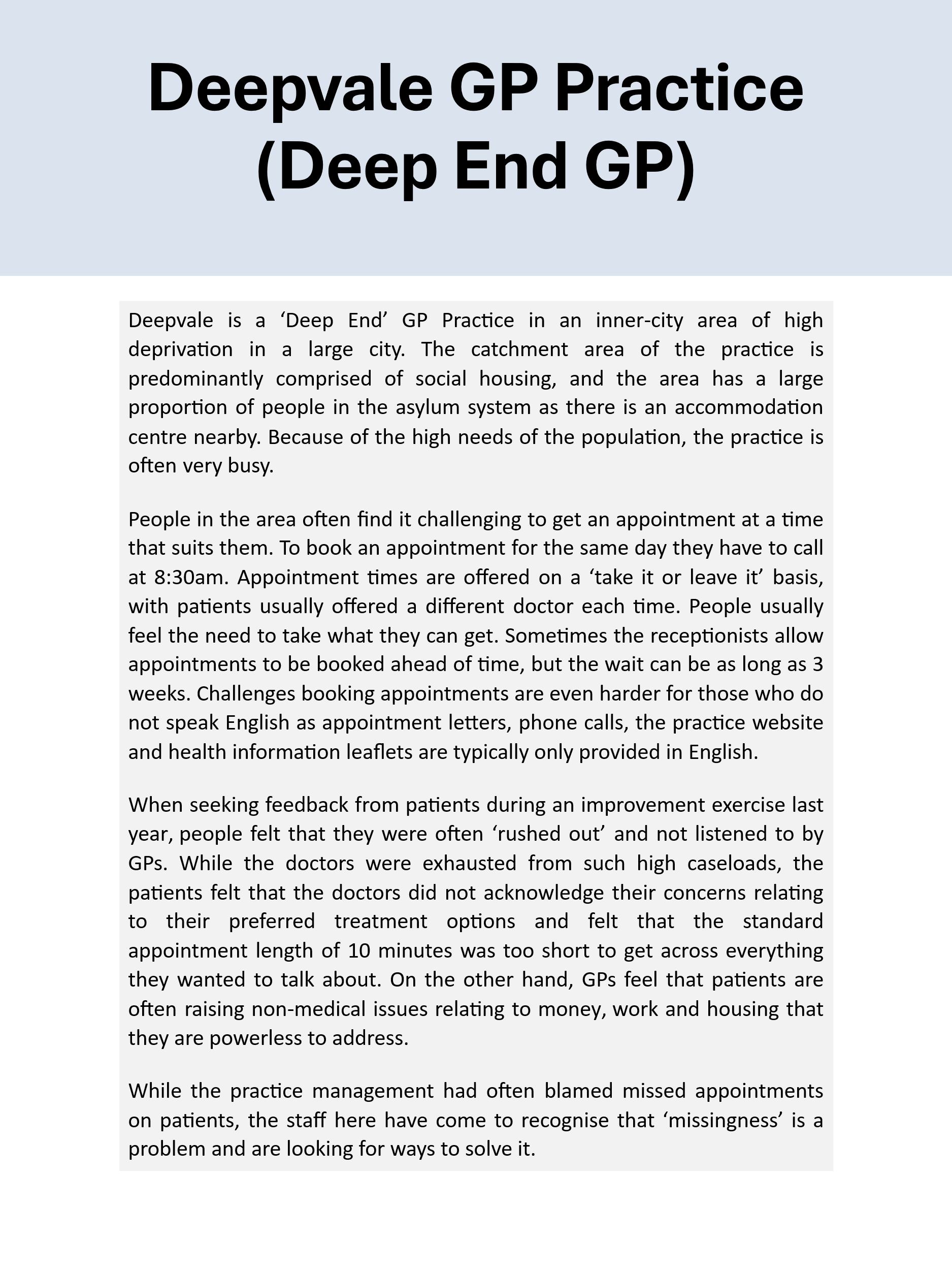

Figure 5 and 6: Template for refined intervention approach for service case study, and example case study of a GP practice

### 7. Contributions of work packages to overall findings

| **Embedding a missingness lens** | | |
| --- | --- | --- |
| **WP1: Realist synthesis** | **WP2: Interviews** | **WP3: Workshops** |
| - Significant support for applying specific resources to carry out additional work with patients in complex circumstances or experiencing access issues. - Promising examples of Quality Improvement, audit, or related approaches (multiple components, locally adapted, seeking to address causes actively, regularly reviewed, including multiple stakeholders). - Staff motivation and knowledge requires resource and support to become meaningful action – improving staff morale and buy-in. - Some evidence of prevailing ‘problematisation’ of missingness in research among staff – but also of motivated staff inhibited by systems and resource-poor environments. - Qualitative research suggests training/education on inclusion health, access issues, mental health, substance use, trauma- and psychologically-informed approaches, relational care and communication. - Training may increase empathy, understanding, motivation and reduce stigma, discrimination – but little concrete evidence or systematic measurement. - Requires time/capacity to attend training; available resources; systems to allow training to be put into practice (supervision, reflective space, protected time, feedback). - Examples combining expertise of patients and staff together in design, implement, review of interventions. - Recruiting staff from specific communities at risk of ‘missingness’ – new perspectives and knowledge; bridging to these communities. - Requires IT infrastructure and data availability to track key indicators, markers and support practice. | - A need to change perspectives, cultures among staff at all levels. - Support for QI approaches – localised, building on staff/patient expertise, reviewed regularly. - A need to closely reflect on how practice dynamics contribute to missingness. - Identified training needs and topics: trauma, stigma, condition competency, health inequalities, missingness. - Training needs to be complemented by resources, and everyday support for staff to implement good practice. - Resourcing is central to incentivising the work and bringing in the capacity and flexibility to carry it out. Current systems do the opposite – inhibit or disincentivise ‘missingness’ work. - Responses to missingness have to be welcoming, non-judgemental, invitational. - A central role for co-designing interventions and services with communities, partner organisations, patients. - No support for punishments, sanctions or coercive approaches to practice – these are stigmatising and exclusionary and disincentivise access. - Practice communications should be accessible, translated and help people understand and engage with primary care. - Hiring from different communities – supporting trust, cultural safety, credibility. - Current systems of targeting, incentivising, resourcing care risk patients being neglected or excluded. Needs to change to encourage and resource this action. - Actively seek feedback and complaints to improve practice. | - A need to change perspectives – to secure ‘buy-in’, ‘commitment’, ‘recognition’ or ‘acknowledgement’ of the issue, and ‘cultural change’ in how to manage it. - Crucial that changes occur at all service levels, among all staff – a consistent, unified approach. - Motivated staff need support; current practice impacted by workloads, limited resources, overwhelming nature of practice. - Staff knowledge as a resource - know who to target, how to change practice, resources required and blockers/barriers. - Major role for training on missingness and its causes, what can be done to address it, and on the needs of specific patient groups. - Major role for training on ‘relational care’ and how to address stigma, exclusion, discrimination, bias, poor communication and work with patients experiencing communication issues. - A role for experts-by-experience in training and awareness-raising. - Increasing staff knowledge needs to be matched by ongoing support – reflective space, supervision, peer support – in an environment conducive to relational care. - Important to have the ‘right’ staff – including hiring from affected communities and reducing ‘distance’ between services and patients. Bringing new perspectives important. - Missingness needs to be measured, monitored and reported transparently, with simple routes to accountability. Suggested a ‘charter’ of commitments. - Requires ongoing monitoring through qualitative and quantitative data, and through conversation within practices. - Essential that practice is resourced/funded/incentivised – or not actively discouraged by systems. |
| **Relationships & Communication** | | |
| **WP1: Realist synthesis** | **WP2: Interviews** | **WP3: Workshops** |
| - Ensuring sufficient staffing and resources for building and maintaining relational care. - In qualitative studies, participants highlight importance on positive relationships and communication, non-judgemental and non-stigmatising practice, continuity, consistency and trust. - Some suggest continuity/relationship brings a sense of reciprocity, loyalty or obligation to attend to protect relationship. - Qualitative research supports addressing power differentials through collaborative, person-centred care acknowledging patients’ broader/structural circumstances. - Qualitative research and quantitative research supports practices of cultural competency and cultural safety (e.g interpreting, staffing changes or ‘matching’, community liaison/outreach, reduced power differentials). - Evidence that relational changes are inhibited by resources (high caseloads, pressure, stress, burnout, vicarious trauma, moral injury). - Qualitative evidence that it is important to reduce repeat/unnecessary disclosure and to avoid excessive gatekeeping activity. | - Training to improve empathy, supportiveness and understanding. - Improving trauma-informed communication practices and responses – building awareness and skills, exploring roots and making changes in practice. - Need to support staff and have systems to support trauma-informed and relational care. - Important to be seen by the ‘right’ clinician – experienced, supportive, trusted, ‘known.’ - Relationships central to other areas – e.g identification (disclosure of sensitive information). - Ensuring language/communication needs are recorded/assessed and used in all contacts. - Contact details and records need to be regularly checked and updated. - Patient-centred, collaborative care gives people a choice, helps secure buy-in and ownership, addresses power imbalances. - Speaking to people/giving information about health and care that is accessible and relevant to their circumstances. - Important for people to have a ‘known person’ to contact for consistency. - Patient-centred care with good communication maximises meaningful and valuable engagements – appointments are worth attending. - Continuity builds trust, enhances disclosure and reduces repeat disclosures, builds loyalty and reciprocity – important for those with traumatic histories. - Continuity does not have to be the same person – about a “consistent response” in a warm, empathetic manner. - For some, continuity is neither wanted nor needed – depends on patient circumstances and preferences. - Providing or supporting access to phones and data to make and maintain contact. | - Both a site for specific actions as and as underpinning the whole intervention. - Importance of empathy, respect, friendliness and human connection in all service interactions. - Patients’ communication needs and preferences should be assessed and adhered to in all service contacts (e.g interpreting, literacy, contact preferences). - Patients should have more options to ‘reach in’ to services to give them information, discuss concerns or care, make changes between or outside of ‘formal’ appointments. - No support for punitive, coercive communication around missed appointments. - Staff training focusing on communication skills and needs e.g trauma-informed practice, relational care/access (see embedding above). - Staff need support, space and time to carry out good relational work, particularly in complex or challenging circumstances. - Importance of continuity and consistency – of practitioners, of approach, of communication style – rooted in principles of patient-centred care. - Addressing power disparities through collaborative, person-centred planning. - Identification-work as relational work; the relationship a precursor for identifying and addressing needs. - Relationships as a form of intervention themselves – building trust with practices and overcoming or avoiding stigma, discrimination, exclusion. |
| **Identification** | | |
| **WP1: Realist synthesis** | **WP2: Interviews** | **WP3: Workshops** |
| - Evidence that current data systems are inadequate for finding and using the data needed. - Different examples of how to define/identify patients within systems (proportion of appointments missed; those with most missed appointments; those with other vulnerability markers e.g multiple health conditions). - Promising studies involve speaking to patients about barriers in order to match activities or provide resources. - Evidence that these contacts have relational benefits: contribute to empathy/understanding from staff and to patients feeling less anxious/more welcome. - Other strategies for barrier-identification include engagement with community groups/organisations. - Multiple suggestions for what should be included in initial/ongoing assessment: patient activation, trauma, Adverse Childhood Experiences, attachment, depression/anxiety, ADHD – but also wider circumstances (holistic/whole person approaches). These are largely untested. - Some qualitative support for patient profile documents – “snapshots” or “passports” to communicate key information. - Qualitative evidence showing that time and good relational practice required to overcome mistrust and past relational difficulties. A relationship may be a pre-requisite to proper identification-work. - Significant evidence that need and circumstances beyond health and healthcare access significantly influence attendance – suggests these need to be addressed within interventions. - Qualitative support for approaches built on patients’ perspectives, expressed needs and priorities. | - Services can take different approaches to identifying ‘high risk’ patients from records – based on health needs or living circumstances, requiring good coding practices and systems. - Strong support for speaking to people sensitively, empathetically and in a trauma-informed way about their barriers to care. - Identification of patients through other practitioners or services, through staff knowledge and experience, or notifications of missed appointments in secondary care. - Identifying and recording access needs and offering *flexibility.* - Strong support for identifying and meeting broader needs and building a broader/deeper model of health – based on patient priorities, collaborative approaches. - Advocating for including support networks (within GDPR and confidentiality rules). - Persistence balanced against the right not to engage – and to understand that this can change. - Computer systems need to be able to ‘flag’ patients regularly for follow up. - Identification should also include strengths, resources – not just problems. - Identification as a part of more robust and comprehensive care planning. - Patient profiles/individual needs reduce repeat disclosures, retraumatisation risks, and should contain agreed actions to facilitate direct access. - Patient profiles/individual needs help create consistency, clear expectations and plans. | - Some support for a ‘phased’ approach – starting with a narrower patient cohort then expanding as resources allow. Narrower cohort could be those at greatest risk. - Staff knowledge of patient cohort will help in identifying who to target. - Practice systems should help identify ‘missing’ patients retrospectively and flag new patients who meet the criteria. - When patients contact practices, systems should flag that they are part of the intervention cohort. - Practice systems/staff should actively code and collect data related to missingness. - Secondary-care ‘missingness’ (e.g missed referral appointments) can be useful for identification. - Where data suggests particular communities are at risk, outreach to those communities for insights. - Contact patients to explore barriers to attendance and their wider health/living circumstances – a holistic, person-centred approach. - Initial contacts should be invitational, welcoming, and is a first step to (re-)establishing a positive relationship – based in curiosity, empathy, showing interest and desire to listen and to help. - Strong support for collaborative, patient-centred “Patient Individual Needs” document – outlining key considerations, commitments and a tailored support plan. Key to consistency and flexible access. - Bringing together staff and patients to discuss causes and possible solutions, as well as other community stakeholders (e.g groups, organisations). - Questions of how to establish boundaries for inclusion/identification given resource considerations. - Temporal elements – need to be able to identify and act when people’s circumstances change. - People may need time to build a relationship before they feel able to disclose. Trust is important. - Some may not wish to be contacted, be part of the intervention, or to continue – persistence balanced against this. |
| **Missingness Coordination** | | |
| **WP1: Realist synthesis** | **WP2: Interviews** | **WP3: Workshops** |
| - Missingness an indirect part of existing research on similar roles - navigators, peer workers, engagement workers, care coordinators, mentors, advocates, coaches, link workers orientation workers – showing positive impacts. - Literature shows that these workers often carry out similar tasks to those outlined here. - Qualitative and quantitative evidence shows the key elements of these roles are: building relationships, developing trust; non-judgemental, empathetic practice; reliability, consistency, responsiveness and flexibility; collaborative, person-centred approaches, tailored approaches; linking, mediating, or bridging between patients and practices or other services; brokering access to wider resources to address wider needs and allow health to become more manageable and a greater priority; flexibility and open-endedness. - Evidence of brokered access activities including acting as a direct point of contact, facilitating rapid/flexible access, advocacy and accompaniment. - Evidence that this work can support patients to develop resources, skills, capacities (confidence, self-efficacy, motivation, self-esteem, expectations for health). - Practice can be compromised when workers are marginalised/peripheral to services, or are ‘co-opted’ into existing practices – need to be able to influence practice change for individual patients and collectively. - Evidence that when this is done, practice can change (better person-centred or relational care, better collaboration). - Peer workers may have benefits – credibility and trust for patients, new perspectives for services, extra levels of ‘safety.’ - Done well, there is evidence that this work also impacts intermediate outcomes related to missingness: patients feeling cared for/understood; reduced delays or barriers to care; improved satisfaction with care; reduced treatment burden; improved coordination; greater understanding of health and health management; increased optimism and quality of life; improved health. - Resources are central: lack of funding can impact holism, flexibility, capacity to build relationships. | - Strong support in general for these approaches, particularly their ‘bridging’ or ‘mediating’ capacity. - Should help address broader needs and social determinants, based on patients’ priorities, and actively support access to resources to address. Can act as a central coordinating point for different types of support. More than just referral but active support to access. - This requires flexible, open-ended, accessible support and inclusive criteria for access. - Coordinators can become main/first point of contact to access care. - Centrality of trust and relationship-building, often overcoming relational injuries – may help rebuild/repair relationships with wider care. - Small caseloads to allow sufficient time and flexibility. - A role for accompaniment to overcome fear, anxiety, stigma, worry, to help communicate needs and ensure care meets needs. - Helping people build confidence, capacity, knowledge, skills, contacts, resources to manage systems in future. - Provided examples of existing approaches to coordination involving many of the tasks here – relationship-building, accessing broader support, accompaniment, advocacy, identification-work, reminders, orientation, follow-up. - Support for peer approaches and hiring from within affected communities – relational benefits as well as practical; modelling change and credibility, power balance, avoiding stigma. - Key qualities: caring, reliable, consistent, assertive, persistent. - Support for hiring those with skills and capacity to manage complexity, risk, safeguarding and emotional demands of the work. - Mixed views on whether this role is an NHS job or should be independent – but need to be sufficiently embedded in practice to discuss cases with care teams, influence practice and systemic change. - Third-sector basis may provide more space for flexibility, creativity. - Coordinators have to be embedded in a wider approach – cannot be constantly fighting the system or be ‘co-opted’ into existing systems or problematic practices. - Primary care as a suitable site for this work – often a first point of contact for help, with access to other parts of the health system, and can hold risk and uncertainty. - Impact may be limited by issues in broader systems/structures – are broader resources available/impactful? - Done well, this may reduce burdens on clinicians or the health service generally. - Paying people well to do this work reflects its importance, difficulty and encourages good practice, retention and quality. | - Strong support for a ‘specialist worker’ focused on missingness. - Relationship-building and ongoing identification-work central to the role – reaching out, creating PIN documents, building ongoing relationships, acting as a point of contact. This projects a sense of care and support. - Should be empowered to bring the patient’s voice and knowledge of their life into the practice and to advocate for changes. - Specific tasks have to be flexible to patients’ individual circumstances and timescales have to be open. - Engaging in ‘bridging’ work – information, navigation, orientation, advocacy, coordinating different forms of care. - Similar work beyond healthcare to address broader needs, competing demands, determinants of ill-health or poor access. - Carrying out contact around appointments and supporting with transport/logistics including accompaniment. - Coaching/mentoring to help develop confidence, skills and self-efficacy. - Embedded in primary care and able to influence system change and flexibility. - Based in primary care because this remains a central and accessible form of help for many patients. - May take some of the complex work away from clinicians and permit them to focus on medical needs while addressing some determinants of those needs. - Missingness may provide an organising principle for similar, pre-existing roles (e.g community links workers). - The levels of consistency, reliability, training and capacity, and the need to embed this work in practice, suggests a paid professional rather than volunteer or non-specific staff member. - Support for different kinds of professional/personal background – a role for expertise by experience but also professional experience in different settings. More important was whether the person could empathise and build relationships and trust. - Importance of choice and consistency for patients in who they work with. - Mixed views on whether this person would be an NHS staff member - which may put off those mistrustful of the service or create a perceived conflict, or a third sector worker – perceived independence but less able to ‘embed’ in practices. - Crucial that this role does not lead to ‘subcontracting’ missingness work to one person – embedding across practice remains important. - Concerns about adding another ‘layer’ of support to those who may already have lots of input. |
| **Flexibility** | | |
| **WP1: Realist synthesis** | **WP2: Interviews** | **WP3: Workshops** |
| - General qualitative support for adjusting appointment-making and attendance systems to patient needs, and tailoring to patients. - Specific support for coordinators acting as brokers of flexible access. - Evidence that offering choice of times may reduce influence of competing demands and maximise convenience. - Qualitative support for offering choice of practitioner(s) to support care continuity. - Support opening appointment times to include evenings and weekends. - Strong qualitative and quantitative evidence for minimising delays to appointments through rapid access, open-access or drop-ins. - Strong qualitative and quantitative support for longer appointments alongside good communication practices – benefits for trust, rapport, comfort, communication. - Strong qualitative support for facilitated access to other services or parts of the healthcare system – reducing delays, overcoming exclusionary eligibilities or punitive responses elsewhere. - Almost no research on home visits beyond qualitative suggestion that they might reduce logistical barriers, health barriers, anxieties/fears. - Outreach care may build links and lead to more regular engagements as well as word-of-mouth to other ‘missing’ patients. - Telehealth/remote care evidence often lacks a missingness lens or the involvement of marginalised patient groups - but evidence that it can reduce missed appointments. Also evidence that missed appointments are stratified along similar lines – many causes remain unaddressed and new barriers evident. - Remote care options may reduce burdens of travel, the influence of competing demands, or reliance on caregivers to travel. - Mixed evidence on relational impacts of telehealth - indications that existing relational dynamics influence this. - Remote care may be particularly appropriate for minor or more transactional appointments. - Evidence that uptake, satisfaction and interest in remote options are lower among groups more likely to be ‘missing,’ including concerns about privacy, safety, confidentiality. - Particular concern about the uniform application of models of remote care without adjustment/design for specific patient groups. - Concerns and evidence around lack of access to devices, credit/data, about complex, inaccessible or unreliable systems, a lack of choice, and lack of safe and private space. - Evidence that uptake and experiences improve with: clear/accessible information on how to use; facilitated access to devices or credit/data; support from coordinators; and staff training. - Remote care needs to be offered as a choice and tailored to needs and preferences – patient-led. | - Particular role for flexibility around different presentations – lateness, unexpected appearances, attendance with friends/family, ‘difficult’ behaviours. - Offering multiple forms of flexibility – how to register, how appointments are made, how/where they are attended, offering choice, longer appointments, wider opening times, telehealth, outreach – tailored to specific patient needs/circumstances. - Identifying and actively offering flexibility has potential to reduce friction and conflict. - Reduce treatment burdens by combining appointments, seeking to maximise value, reducing unnecessary attendances (LTC) - Offering drop-ins, same-day or rapid access to specific patients has relational and practical benefits. Concern that general drop-ins would be hard to manage. - Strong advocacy for offering home visits to build relationships, secure buy-in. Requires resources. - Outreach and in-reach can build connections to healthcare for those most included, particularly with consistency over time. - Longer appointments increase the value of those appointments, supporting exploration of complexity and good communication practice. - Telehealth may be suitable under some circumstances – may permit longer appointments, reduce resource requirements or burdens of attendance. Tailoring and choice are central. - Carrying out care at home or in other spaces builds insight into patients’ lives, circumstances. - Facilitating rapid/flexible access to other aspects of the health system (e.g secondary/specialist care). - Flexibility is not limitless – boundaries are important for consistency and clarity, within a relational approach. - Service co-location, coordination or integration to maximise access to different forms of care with minimal fragmentation or resource requirements. - Maximising those appointments that do happen – do what you can, when you can. - Coordinated care approaches to reduce burdens/unnecessary care interactions. | - Important to offer walk-in or drop-in availability to specific patients (or during specific time periods). - Support for increasing opening hours (evenings, weekends) and reserving for specific patients to avoid ‘capture’ by others. - Offering choice/control over: when to attend, how to attend (e.g in-person or remote), how to be contacted, and who they might see. - Offers of flexibility in how appointments are made – in-person, online, via coordinators**.** - Offering longer appointments allows extra time for communication, discussion, negotiation, complexity and may allow people to feel more comfortable. - Offering flexibility demonstrates a willingness to change care and support access. - Flexibility offers should include changes in the site of care including home visits, outreach/inreach, and remote care options – determined by patient needs/circumstances. - Home visits: patients may feel safer, may be unable to travel for health, financial, time reasons, and professionals may be able to get insight into living circumstances. - Outreach: into key spaces where ‘missing’ patients are likely to be. - Remote care: may be appropriate under some circumstances; support patients to feel safe/secure; and reduce the burdens of travel. Concerns about tech access, affordability, reliability, and whether these are designed to the benefit of services and not patients. Concerns too about relational impacts. Coordinators could play a supporting role. - In all flexibility, choice and design via a missingness lens is central. - Flexibility extends belong primary care to secondary/specialist care – patients should be prioritised for rapid, facilitated/supported access. - Support for co-located, integrated services and other activities bringing services together to reduce burdens of multiple ‘candidacies’ for patients. Activities include multi-service appointments; hub models, coordinated care planning, and bringing services into primary care settings. |
| **Transport and Logistics** | | |
| **WP1: Realist synthesis** | **WP2: Interviews** | **WP3: Workshops** |
| - Mixed evidence on transport interventions – reflecting weaknesses in design or delivery and an excessive focus just on logistical causes. - Qualitative evidence that transport systems can be inaccessible, complex, confusing and exclusionary. - Existing transport systems often require more notice to arrange than patients have. - Some qualitative and quantitative support for pre-emptively providing travel tickets, vouchers or permits, or arranging taxi transport. - Almost no research on home visits beyond qualitative recommendations that they might reduce logistical barriers, health barriers, anxieties/fears and be more comfortable. - Coordinator roles often identify and support transport, including accompaniment to support navigation and overcome potentially difficult or challenging journeys. - Accompaniment also has relational benefits within appointments – supporting good communication, advocacy work, and increasing a sense of safety. - Trauma-informed or psychologically-informed waiting rooms and spaces. - Co-located services – may bring more ‘missing’ patients into spaces where primary care services are and may facilitate access. | - A particular role for accompaniment to overcome fear, anxiety, stigma, worry and to support communication. - Eligibility for transport help needs to be broad, equitable and accessibly designed. - Systems for access need to be accessible and involve minimal effort/resources. - A role for delivering care in other settings through outreach.   • Changing physical practice environments to feel comfortable, welcoming, inclusive and safe.  • Co-locating services in ‘hub’ models to minimise resource burdens of moving between spaces/services. | - Described a range of possible activities on a spectrum, to be offered depending on circumstances or assessment of need. - Reimbursing costs suggested – but systems are often complicated and leave people out-of-pocket. - Pre-emptively sending tickets for different forms of travel suggested but concerns about how to get tickets to people and possible tech barriers. - Taxi services suggested but concerns about costs for services. - Strong support for coordinators accompanying people on their journeys for safety, reassurance and practical navigation support. - Other alternatives – home visits, outreach to other settings, remote care – discussed in flexibility. - Systems for accessing this help need to be publicised or offered to patients and easy to access or arrange. - Making changes to physical spaces – making them calmer, quieter, more welcoming, trauma-informed and with private spaces. - Support for co-located, integrated services and other activities bringing services together to reduce burdens of multiple ‘candidacies’ for patients. Activities include multi-service appointments; hub models, coordinated care planning, and bringing services into primary care settings. |
| **Contact Around Appointments** | | |
| **WP1: Realist synthesis** | **WP2: Interviews** | **WP3: Workshops** |
| - Excessive focus on ‘simple’ reminders in the evidence base – rarely focused on missingness or applied/designed with an equity/inclusivity lens. - Evidence that reminders can reduce missed appointments generally, and that ‘stepped’ or repeat reminders may be more beneficial as they support both planning and memory. - Evidence that they are less effective (and potentially ineffective) among marginalised patient groups or those experiencing greatest barriers. - Concerns about access to phones or technology, data/credit, changing contact details or circumstances impacting effectiveness. - Qualitative support that there is a role for reminders and that patients want or appreciate them. - Evidence suggests a good reminder system requires: Work to address inequities in patient contact data; tailoring to fit patient contact preferences (methods, language, frequency, suitability); addressing/respecting concerns about privacy, security, confidentiality; clear information about the appointment; the means to respond the service. - No evidence linking ‘nudge’ reminders to missingness outcomes. - Limited evaluative evidence for personalised reminders but support for contact to build relationships; provide orientation, reassurance and practical support; discuss concerns or anxieties. - Contact should not be coercive or exclusionary. - Support for proactive/assertive outreach for ongoing ‘missingness.’ - Qualitative support for systems allowing patients to make more meaningful, direct contact with services – to provide information, seek advice or support. Coordinators may be central to this. | - Low effort approaches (SMS, letters, calls) may be missed, not received, ignored or not trusted, may feel tokenistic or exclusionary. - Reminders alone are insufficient to address complex causes. - Strong support for personalised approaches to contact based on good relational principles. - Patients given choice in how to be contacted, when, by whom – tailoring. - Contact needs to be expected, otherwise risks being ignored (e.g private numbers). - Contact involving orientation, exploring barriers, checking in, reminding. - Needs to be invitational, positive, non-judgemental and supportive. - Persistence and ‘stickability’ needs to be balanced against the right not to engage – and to understand that this can change. - Advocating for involvement of support networks (within GDPR and confidentiality rules). - Following up on missed appointments – assertive but not exclusionary. - Patients also need the means to easily contact services to cancel, rearrange, discuss appointments. - ‘Nudge’ approaches are unhelpful, unwelcome, lack empathy and may deter access. | - A role for basic reminders – written, with details of the appointment to avoid confusion or anxiety (where, when, who with, what service). - Support for sending multiple reminders by different means/methods – more likely to be received and to support travel arrangements and remembering. - Strong support for personalised pre-appointment contact – discuss concerns, arrange plans to attend, orientation and information, contribute to relationship-building. Carried out by coordinators. - Strong support for follow-up calls by coordinators to check in, demonstrate care, explore barriers. Part of ongoing identification and relationship-building. - Design of any reminder system needs to occur with a missingness lens to avoid worsening gaps. - Concerns about lack of a ‘missingness’ lens in reminding contributing to gaps – patients may avoid unknown numbers or unsolicited contact; issues of tech access and literacy or access to data/credit. - Reminders, while useful, still leave a lot of causes untouched. - A need to fit the contact strategy to patients circumstances, needs, preferences as noted in the PIN. - A role for contacting significant others with patient permission. - Requires work to ensure contact details and preferences are up-to-date. |
| **Other interventions** | | |
| **WP1: Realist synthesis** | **WP2: Interviews** | **WP3: Workshops** |
| - Some papers discussing incentives (financial, in-kind e.g food, contingency management) but without a missingness lens and evidence that they can sustain problematic power dynamics and undermine trust and collaboration. - Some methods of scheduling or booking (including overbooking) discussed but of benefit to services and actively disadvantaging ‘missing’ patients. | - Some things need to change beyond the healthcare system – structural dynamics (poverty, housing, health determinants, asylum system) as well as political/policy conversations around healthcare use and health inequalities. - Incentivising attendance **–** providing basic essentials, even financial incentives – suggested occasionally but not central to narratives. | - No support for punitive, sanctioning or coercive approaches, including ‘nudge’ interventions. - No support for ‘overbooking’ or scheduling interventions aimed at benefiting practices and not patients. |

1.	Duncan E, Cathain A, Rousseau N, Croot L, Sworn K, Turner KM, et al. Guidance for reporting intervention development studies in health research (GUIDED): an evidence-based consensus study. BMJ Open. 2020;10(4):e033516.
